# Large scale brain network dynamics in very preterm children and relationship with socio-emotional outcomes

**DOI:** 10.1101/2022.03.18.22272505

**Authors:** Vanessa Siffredi, Maria Chiara Liverani, Lorena G. A. Freitas, D. Tadros, Y. Farouj, Cristina Borradori Tolsa, Dimitri Van De Ville, Petra Susan Hüppi, Russia Hà-Vinh Leuchter

## Abstract

Children born very preterm (VPT; < 32 completed weeks of gestation) are at high risk of neurodevelopmental and neurobehavioral difficulties associated with atypical brain maturation. The analysis of large-scale brain network dynamics during rest allows to investigate brain functional connectivity and their association with behavioural outcomes. Of interest, prematurity has been associated with atypical socio-emotional development with significant implications for the forming of peer relationships, adaptive functioning, academic achievement and mental health. In this study, we extracted dynamic functional connectivity by using the innovation-driven co-activation patterns (iCAPs) framework in preterm and full-term children aged 6 to 9 to explore changes in spatial organisation, laterality and temporal dynamics of spontaneous large-scale brain activity. Multivariate pattern analysis was used to explore potential biomarkers for socio-emotional difficulties in preterm children. Results show a spatial organisation of 13 networks retrieved comparable to full-term controls. Dynamic features and lateralisation of network brain activity were also comparable across groups for all large-scale brain networks. Despite apparent similarities in terms of dynamical functional connectivity parameters, multivariate pattern analysis unveiled group differences in their associations with socio-emotional abilities. While a pattern of decreased engagement in certain brain networks were associated with better socio-emotional abilities in full-term controls; in the VPT group, better socio-emotional abilities were associated with coordination of activity across different networks, i.e., coupling duration between different pairs of networks. It is possible that group differences reflect reduced degree of maturation of functional architecture in the VPT group for socio-emotional abilities.

## Introduction

Preterm birth occurs during key phases of interrelated neurobiological processes underlying brain development. This seem particularly relevant in infants born very preterm (VPT; < 32 completed weeks of gestation). As a consequence, VPT children are at high risk of neurodevelopmental and neurobehavioral difficulties associated with atypical brain structural and functional maturation (Volpe, 2009).

Magnetic resonance imaging (MRI) techniques provide powerful, non-invasive tools to delineate aberrant brain development related to prematurity. In particular, resting state-functional MRI (rs-fMRI) has emerged as a promising tool for studying neural networks underlying typical and atypical brain development (Ellard et al., 2021). Functional connectivity (FC) derived from rs-fMRI is defined as the temporal dependence of neuronal activity patterns of anatomically separated brain regions (Fox and Raichle, 2007; Van Den Heuvel and Pol, 2010). FC between regions in the absence of goal-directed activity and stimulation are used to identify networks with synchronous, spontaneous neuronal activity, termed resting state networks (Fox and Raichle, 2007; Van Den Heuvel and Pol, 2010). The functional relevance of resting state networks has been inferred not only from their spatial overlap with regions known to underpin sensorimotor and cognitive functions but also from their overlap with known white-matter anatomical pathways (Biswal et al., 1995; Fox et al., 2005; Greicius et al., 2009; Skudlarski et al., 2008).

To date, most studies on rs-fMRI in VPT individuals of different ages from newborn to adolescent have used static FC, i.e., the correlation between the activation in different brain regions over the whole scanning time (Preti et al., 2017). Using a seed based correlation approach - in which few region of interest (ROI) time series are selected a priori and voxelwise cross-correlation is computed across the whole brain (Fox et al., 2005) - studies in VPT individuals at different ages have found altered FC in various brain networks using ROIs such as superior temporal, amygdala, posteromedial and lateral parietal, prefrontal or lateral and superior sensorimotor regions (Carter et al., 2004; Johns et al., 2019; Papini et al., 2016; Wehrle et al., 2018; Wilke et al., 2014). Similarly, alterations of different resting state networks have been found in VPT individuals when using whole-brain data-driven approaches, including FC of the salience, default mode or frontal networks (Ball et al., 2016; Damaraju et al., 2010; Lordier et al., 2019; White et al., 2014). However, recent studies suggest that stationary measures might be too simplistic to capture the full extent of resting state activity as that they ignore the inherently dynamic nature of brain FC (Christoff et al., 2016; Karahanoğlu and Van De Ville, 2017; Preti et al., 2017). In this perspective, dynamic approaches have been developed with the potential to identify meaningful variations over time in FC between different brain regions.

Among various methods to identify dynamic FC, the innovation-driven co-activation patterns (iCAPs) framework detects moments of significantly changing brain activity to extract large-scale brain networks and their dynamic properties (Karahanoğlu and Van De Ville, 2015; Karahanoğlu and Van De Ville, 2017; Zöller et al., 2018). Using a whole-brain data-driven approach, this framework not only allows to extract spatial and temporal characteristics of large scale brain networks but also temporal overlaps between these different networks. Using the iCAPs framework, recent studies conducted in children, adolescents and adults have found significant association between large scale brain dynamics and psychiatric symptoms including depression, anxiety and psychotic symptoms (Piguet et al., 2021; Zöller et al., 2019).

In VPT children, studies show that approximately 25% of them experience behavioural problems (Arpi and Ferrari, 2013; Johnson and Marlow, 2011). Of particular interest, socio-emotional abilities have been found to have important implications for the forming of peer relationships, adaptive functioning, academic achievement and mental health (Izard, 2011; Montagna and Nosarti, 2016a; Reyes et al., 2021). Prematurity has been associated with atypical socio-emotional development as early as the first year of life and extend from difficulties in emotional information processing, social understanding, emotion regulation, socialising, peer relationship as well as internalising problems (Bhutta et al., 2002; Healy et al., 2013; Hille et al., 2001; Johnson et al., 2015; Johnson et al., 2010; Jones et al., 2013; Landry et al., 1990; Langerock et al., 2013; Montagna and Nosarti, 2016b; Treyvaud et al., 2013; Witt et al., 2014). Importantly, deficits in socio-emotional processing and regulation in early life are considered precursors of later psychiatric and mental health problem (Briggs-Gowan and Carter, 2008; Carter et al., 2004). In VPT individuals, these difficulties have indeed been found to be long-lasting, with consequences observed in social, occupational and family functioning through adolescence into adulthood (Mendonça et al., 2019; Montagna and Nosarti, 2016b; Saigal et al., 2016); as well as increased risk of developing psychiatric disorders as adults, including depression, bipolar affective disorder, anxiety disorder and schizophrenia (Nosarti et al., 2012; Räikkönen et al., 2008; Walshe et al., 2008). In line with these studies, alteration in structural brain architecture has been associated with atypical socio-emotional development in VPT children (Fischi-Gomez et al., 2015; Kanel et al., 2021). Consequently, understanding brain functional underpinning of socio-emotional abilities in young VPT children might therefore inform on potential biomarker of socio-emotional difficulties and vulnerability of future psychiatric disorders.

In this study, we extracted dynamic FC by using the iCAPs framework in young preterm and full-term children aged 6 to 9 to explore changes in spatial organisation, laterality and temporal dynamics of spontaneous large-scale brain activity. Multivariate pattern analysis was used to explore potential biomarkers for socio-emotional difficulties in preterm children.

## Methods and Materials

### Participants

227 VPT children born before 32 gestational weeks between 01.01.2008 and 01.05.2013, in the Neonatal Unit at the Geneva University Hospital (Switzerland) and followed up at the Division of Child Development and Growth, were invited to participate in the “Via-à-vis interventional study” between January 2017 and July 2019. VPT children were excluded if they had an intelligence quotient below 70, sensory or physical disabilities (cerebral palsy, blindness, hearing loss), or an insufficient understanding of French. A total of 45 VPT participants aged between 6 to 9-year-old were enrolled. Moreover, 17 term-born controls aged between 6 to 9-year-old were recruited through the community.

Described in details below, of the 62 enrolled participants, 9 participants were excluded as they did not complete both the brain MRI scan and the neuropsychological assessment (VPT, n=7; full-term controls, n=2); and 13 participants were excluded due to high level of motion artefacts in rs-fMRI sequence described in the previous section (VPT, n=10; full-term controls, n=3). The final sample included 40 participants between 6 and 9 years of age: 28 VPT and 12 full-term participants.

This study was approved by the Swiss Ethics Committees on research involving humans, ID: 2015-00175. Written informed consent was obtained from the principal caregiver and from the participant.

### Neuropsychological measures

The Kaufman Assessment Battery for Children – 2nd Edition (K-ABC-II; (Kaufman and Kaufman, 2013)) was used to evaluate the Fluid-Crystallized Index (FCI) as a measure of general intellectual functioning. For children younger than 7 years of age, the FCI is derived from a linear combination of 11 core subtests; and for older children, it is derived from 10 core subtests. The FCI have a mean of 100 and a standard deviation of 15.

Participants’ socio-emotional outcomes was assessed using four different measures. First, subtests of the Developmental Neuropsychological Assessment - 2nd Edition (NEPSY-II (Korkman et al., 2007)) including: a) the Affect Recognition subtest giving a total score assessing facial emotional recognition; and b) the Theory of Mind subtest giving a total score measuring the ability to understand mental contents, such as belief, intention or deception. As the Theory of Mind subtest does not provide a standard score, raw scores for both measures of the NEPSY-II were regressed on age at testing and standardised residuals was used as a score, called affect recognition and theory of mind. Second, the Internalised Score of the Strength and Difficulties Questionnaire – parent version (SDQ) was used to assess emotional and peer problems in daily life (Goodman, 1997, 2001). It rates participant’s internalised difficulties over the previous 6 months. The Internalised Score of the SDQ is scored on a Likert scale and is the sum of the emotional and peer problems scales. As standardised scores are not available for this measure, raw scores were regressed on age at testing and standardised residuals was used as a score, called internalised problems. Higher internalised scores reflect increased internalised difficulties in daily life. Finally, the Emotional Control Scale of the Behaviour Rating Inventory of Executive Function, parent version (BRIEF (Gioia et al. (2000)) was used to measure the extent to which the child is able to mediate emotional responses in daily life. As standardised scores are available for this measure, standardised scores were used (mean = 50, SD = 10), called emotional control. Higher emotional control scores reflect increased difficulties in emotional control.

### Magnetic Resonance Imaging (MRI) acquisition and preprocessing

#### Magnetic Resonance Imaging acquisition

MRI data were acquired at the Campus Biotech in Geneva, Switzerland, using a Siemens 3T Magnetom Prisma scanner. All participants completed a simulated “mock” MRI session prior to their MRI scan. This preparation process was conducted by trained research staff and allowed participants to familiarise themselves with the scanner and the scanning process, eventually raising any concerns they might have had prior to the MRI scan. Furthermore, this process is known to facilitated acquisition of good quality MRI images in children (de Bie et al., 2010; Tamnes et al., 2018). Structural T1-weighted MP-RAGE (magnetization-prepared rapid gradient-echo) sequences was acquired using the following parameters: voxel size = 0.9 × 0.9 × 0.9 mm; repetition time (TR) = 2,300 ms; echo time (TE) = 2.32 ms; inversion time (TI) = 900 ms; flip angle (FA) = 8°; and field of view (Fov) = 240 mm. Resting state functional images were T2*-weighted with a multislice gradient-echo-planar imaging (EPI) sequence of 64 slices; voxel size = 2 × 2 × 2 mm; TR = 720 ms; TE = 33 ms; Fov = 208 mm. During the rs-fMRI sequence, children were asked to keep their eyes closed and engage in mind wandering. Finally, a fieldmap was acquired each time a participant entered the scanner, with TR = 627 ms; TE1 = 5.19 ms; TE2 = 7.65 ms; and FA = 60°.

#### Resting-State functional MRI data preprocessing

Our data were preprocessed using SPM12 (Wellcome Department of Imaging Neuroscience, UCL, UK) in MATLAB R2016a (The MathWorks, Inc., Natick, Massachusetts, United States). Rs-fMRI data were converted from the native DICOM to NIFTI format and the preprocessing pipeline described by Freitas, Liverani and colleagues was used (Freitas et al., 2021). Rs-fMRI images were spatially realigned and unwarped, respectively, to correct for motion artefacts and potential geometric distortions. The unwarping step allows to improve the co-registration between structural and functional images and to reduce the distortion variability across subjects during spatial normalisation to a common space (Hutton et al., 2002). Functional images were then coregistered to their corresponding structural images in subject space and smoothed with a Gaussian filter of full width at half maximum (FWHM) = 6 mm. Structural images were segmented with the SPM12 segmentation algorithm to automatically identify different tissue types within the images, i.e., grey matter, white matter, cerebrospinal fluid (Ashburner and Friston, 2005); and a study-specific template was generated using Diffeomorphic Anatomical Registration using Exponential Lie algebra (DARTEL (Ashburner, 2007)) that will be used in the Innovation-Driven Co-Activation Patterns (iCAPs) framework described below. Finally, the first five rs-fMRI images were excluded, and average white matter and cerebrospinal fluid signals were regressed out from the BOLD time series.

#### Head motion

For rs-fMRI data, the mean framewise displacement for each frame was computed to quantify the extent of head motion from volume to volume for each participant (Power et al., 2012; Power et al., 2014). Following Power and colleagues’ recommendations, volume censoring (“scrubbing”) for motion correction were applied to frames with a mean framewise displacement above 0.5 mm, as well as one frame before and two after. Moreover, if more than 20% of the frames had to be scrubbed, the participant was removed from further analyses. Based on these criteria, 10 VPT and 3 full-term control participants were excluded from further analyses.

Innovation-Driven Co-Activation Patterns (iCAPs) and extraction of iCAPs activation measures

#### Innovation-Driven Co-Activation Patterns (iCAPs)

The innovation-driven-co-activation-pattern (iCAP) analysis is a novel state-of-the-art rs-fMRI analysis tool that allows to derive a set of whole-brain spatial patterns of regions whose activity simultaneously increases or decreases, thus characterised by similar functional dynamic behaviour (Karahanoğlu and Van De Ville, 2015). For a comprehensive explanation of the methodology and procedure, please refer to Karahanoglu and colleagues (2015) and to Zöller and colleague (2019). We tailored the openly available MATLAB code (https://c4science.ch/source/iCAPs/) MATLAB vR2016a (The MathWorks, Inc., Natick, MA) to apply the iCAPs framework in VPT and full-term participants. The overall routine is composed of 4 steps:

- Total activation (TA): In this first step, we employed TA which applies a voxel-wise hemodynamically-informed deconvolution (Farouj et al., 2017; Karahanoğlu et al., 2013) to the fMRI timeseries in a way that promotes the rareness of activity transients and spatially coherent activations. TA provides three types of information: (1) activity-related signals that are denoised fMRI signals; (2) sustained, or block-type, activity-inducing signals that are deconvolved signals; (3) innovation signals that are the derivative of the activity-inducing signals and encode transient brain activity episodes as spikes.
- Significant transients detection: *Innovation* signals are computed as the temporal derivative of the deconvolved signals. The obtained signals can be seen as a representation in terms of transients in neural activity, where large amplitude transients implicitly identify change-points. Significant transients were determined using a two-step thresholding procedure. A temporal threshold estimated from a surrogate distribution that keeps only transient larger than 95% or lower than 5%. Then, a spatial thresholding procedure was applied, in which a frame was considered significant if at least 5% of the grey matter voxels were active. The frames showing significant transients are called *innovation* frames, and allow to identify time-points when a given region in the brain undergoes an increase or decrease in activity.
- Aggregation: The significant, i.e. *innovation*, frames were warped into MNI (Montreal Neurologic Institute) space via a study-specific DARTEL template previously created (see preprocessing). All frames were then aggregated for clustering.
- Clustering: The retained *innovation* frames, underwent K-means clustering to obtain simultaneously transitioning brain patterns, i.e., the iCAPs. The optimum number of 13 clusters was determined by consensus clustering and following recommendation of previous studies (Karahanoğlu et al., 2013; Monti et al., 2003; Zöller et al., 2019) (Supplementary Figure S1 & S2).
- Time courses extraction: Time courses were obtained for all iCAPs using spatiotemporal transient-informed regression (Zöller et al., 2018).

#### Extraction of temporal properties

For computation of temporal properties, iCAPs’ time series were recovered by backprojecting each iCAP into subjects’ activity-inducing signals; i.e., block-type activity representations recovered by TA. For each iCAP, we then computed two measures representing the temporal characterisation of iCAPs: 1) occurrence, i.e., the number of activation blocks, 2) total duration, i.e., the total duration of overall activation as percentage of the total nonmotion scanning time.

To explore the dynamic interactions during resting state, coupling and anticoupling duration of each pair of iCAPs were calculated as time points of same-signed or oppositely signed coactivation measured as percentage of the total nonmotion scanning time or as Jaccard score; i.e., percent joint activation time of the two respective iCAPs.

#### Extraction of laterality measure

To explore the laterality of brain networks, the iCAPs maps were co-registered MNI symmetrical template, available at http://www.bic.mni.mcgill.ca/ServicesAtlases/ICBM152NLin2009. Based on previous studies, the laterality of activity maps aimed at exploring possible asymmetry effect between the two hemispheres by comparing lateralised amplitude maps of the iCAPs (Karolis et al., 2019). Therefore, the amplitudes of these patterns reflect the mean activity amplitude for each voxel when contributing to a certain network. In order to obtain an Amplitude Laterality Index (A-LI) for each voxel, we flipped the left hemisphere maps and subtracted them from unflipped right hemisphere maps (Siffredi et al., 2021). Positive and negative values in these A-LI maps reflect, respectively, right and left lateralisation. These maps were then averaged for each iCAPs in order to obtain one A-LI for each iCAPs and each participant.

### Statistical analyses

#### Group comparisons of iCAPs activation measures

Measures of occurrence and total duration of each iCAPs as well as coupling and anticoupling between each pair of iCAPs were compared between the VPT and full-term groups two-sample t-tests. Similarly, A-LI laterality measure was compared between the VPT and full-term groups using two-sample t-tests. The p values were corrected for multiple comparisons with the false discovery rate (FDR, (Benjamini and Hochberg, 1995)). Analyses were performed using the R software version 4.0.3, and R studio version 1.3.1093 (Team, 2020; Team, 2013).

#### Multivariate correlation between iCAPs’ time courses and socio-emotional measures

To evaluate multivariate patterns of correlation between iCAPs temporal characteristic and socio-emotional measures, we used partial least squares correlation (PLSC). A publicly available PLSC implementation in MATLAB was used: https://github.com/danizoeller/myPLS (Kebets et al., 2019; Zoller et al., 2019). PLSC is a data-driven multivariate technique that maximizes the covariance between two matrices by identifying latent components which are linear combinations of the two matrices, i.e., socio-emotional and iCAPs temporal characteristics measures (McIntosh and Lobaugh, 2004).

For the socio-emotional scores, the 4 scores of affect recognition, theory of mind, internalised problems and emotional control were considered. For affect recognition, theory of mind and internalised problems, raw scores were regressed on age at testing and socio-economic status (i.e., Largo score) confounds, and the standardised residuals were used for each score. For emotional control, standard scores were regressed on socio-economic status (i.e., Largo score) and the standardised residuals were used. Socio-emotional scores were stored in a 40 × 4 matrix denoted X, in which each row represents one participant and the matrix’s 4 columns consist of the 4 socio-emotional scores considered. For each iCAPs temporal characteristics, PLSC was repeated four times, each time considering only one temporal characteristics, i.e., occurrence, total duration, coupling, anticoupling. First, occurrence measures were gathered in a 40 × 13 matrix denoted Y, with each row matching one participant and each column representing the occurrence of each of the 13 iCAPs. Second, total duration measures were gathered in a 40 × 13 matrix denoted Y, with each row matching one participant and each column representing the total duration of each of the 13 iCAPs. Third, coupling measures were stored in a 40 × 78 matrix denoted Y, in which each row represents one participant and the matrix’s 78 columns consists of the coupling duration for each pair of iCAPs. Finally, anticoupling measures were stored in a 40 × 78 matrix denoted Y, in which each row represents one participant and the matrix’s 78 columns consists of the anticoupling duration for each pair of iCAPs. A cross-covariance matrix was then computed between X (participants x socio-emotional scores) and Y (participants x iCAPs temporal characteristics). Singular value decomposition was then applied to this cross-covariance matrix, resulting in latent components. Statistical significance of multivariate correlation patterns, i.e., latent component, was assessed with permutation testing (1000 permutations) and considered robust at p<0.01 following guidelines of previous studies (McIntosh and Lobaugh, 2004; Zöller et al., 2019). Stability of saliences were estimated using bootstrapping (500 bootstrap samples with replacement). Bootstrap ratio z-scores for each socio-emotional measure and iCAPs temporal characteristics were obtained by dividing each socio-emotional and iCAPs temporal characteristics weight by its bootstrap-estimated standard deviation, and a p-value was obtained for each bootstrap ratio z-score. The contribution of socio-emotional and iCAPs temporal characteristics weights for a given latent component was considered robust at p < 0.01 (i.e., absolute bootstrap ratio z-scores above 3 or below −3).

## Results

### Participant characteristics

Of the 62 participants enrolled, 9 participants were excluded as they did not complete both the brain MRI scan and the neuropsychological assessment (VPT, n=7; FT, n=2); and 13 participants were excluded due to high level of motion artefacts in rs-fMRI sequence described in the previous section (VPT, n=10; FT, n=3). The final sample included 28 VPT and 12 full-term participants between 6 and 9 years of age (Table 1). Baseline characteristics were comparable between VPT and full-term participants for sex, age at assessment and the Fluid-Crystallized Index. Socio-economic status, as measured by the Largo score (Largo et al., 1989), showed significant group difference, with lower socio-economic status (i.e., higher Largo score) in the VPT group compared to the full-term group. Socio-emotional outcomes were comparable between the VPT and full-term groups across all scores, i.e., affect recognition, theory of mind, internalised problems and emotional control scores.

**Table 1.**
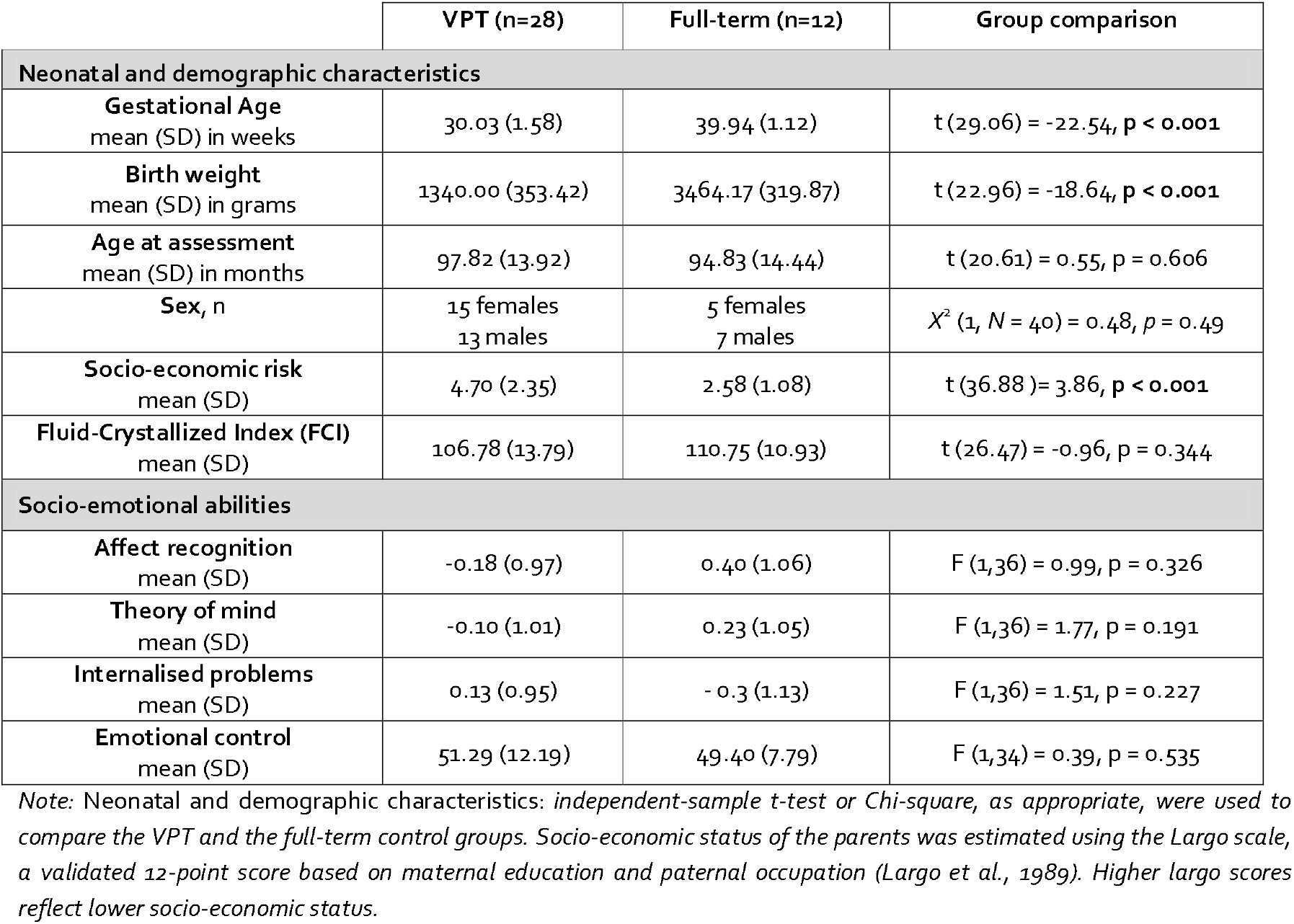
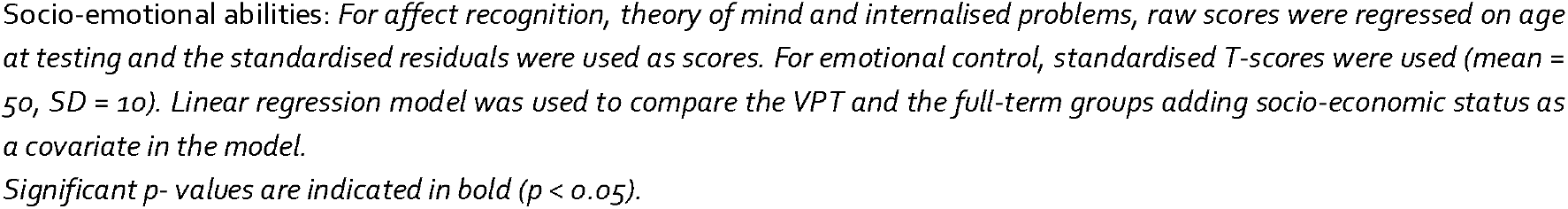
Neonatal and demographic characteristics, as well as socio-emotional outcomes of the VPT and full-term participants

|  | VPT (n=28) | Full-term (n=12) | Group comparison |
| --- | --- | --- | --- |
| **Neonatal and demographic characteristics** |  |  |  |
| **Gestational Age**<br>mean (SD) in weeks | 30.03 (1.58) | 39.94 (1.12) | $t(29.06) = -22.54, p < 0.001$ |
| **Birth weight**<br>mean (SD) in grams | 1340.00 (353.42) | 3464.17 (319.87) | $t(22.96) = -18.64, p < 0.001$ |
| **Age at assessment**<br>mean (SD) in months | 97.82 (13.92) | 94.83 (14.44) | $t(20.61) = 0.55, p = 0.606$ |
| **Sex, n** | 15 females<br>13 males | 5 females<br>7 males | $\chi^2(1, N = 40) = 0.48, p = 0.49$ |
| **Socio-economic risk**<br>mean (SD) | 4.70 (2.35) | 2.58 (1.08) | $t(36.88) = 3.86, p < 0.001$ |
| **Fluid-Crystallized Index (FCI)**<br>mean (SD) | 106.78 (13.79) | 110.75 (10.93) | $t(26.47) = -0.96, p = 0.344$ |
| **Socio-emotional abilities** |  |  |  |
| **Affect recognition**<br>mean (SD) | -0.18 (0.97) | 0.40 (1.06) | $F(1,36) = 0.99, p = 0.326$ |
| **Theory of mind**<br>mean (SD) | -0.10 (1.01) | 0.23 (1.05) | $F(1,36) = 1.77, p = 0.191$ |
| **Internalised problems**<br>mean (SD) | 0.13 (0.95) | -0.3 (1.13) | $F(1,36) = 1.51, p = 0.227$ |
| **Emotional control**<br>mean (SD) | 51.29 (12.19) | 49.40 (7.79) | $F(1,34) = 0.39, p = 0.535$ |

*Note:* Neonatal and demographic characteristics: independent-sample $t$ -test or Chi-square, as appropriate, were used to compare the VPT and the full-term control groups. Socio-economic status of the parents was estimated using the Largo scale, a validated 12-point score based on maternal education and paternal occupation (Largo et al., 1989). Higher largo scores reflect lower socio-economic status.
Socio-emotional abilities: *For affect recognition, theory of mind and internalised problems, raw scores were regressed on age at testing and the standardised residuals were used as scores. For emotional control, standardised T-scores were used (mean = 50, SD = 10). Linear regression model was used to compare the VPT and the full-term groups adding socio-economic status as a covariate in the model.*
*Significant p-values are indicated in bold ( $p < 0.05$ ).*

### Extracted innovation-driven co-activation patterns (iCAPs)

The iCAPs framework was applied to rs-fMRI scans of both VPT and full-term participants. The 13 extracted spatial maps represented in each iCAP correspond to well-known resting state networks and were reminiscent of common task-related and cognitive networks typically observed in fMRI studies (Van Den Heuvel and Pol, 2010). They also correspond to iCAP networks identified in previous studies (Bommarito et al., 2021; Piguet et al., 2021; Siffredi et al., 2021; Zöller et al., 2019). Specifically, the obtained networks included sensory-related networks, i.e., sensori-motor/auditory, primary and secondary visual; higher level cognitive network, i.e., the fronto-parietal, frontal and fronto-temporal left and right networks; and default-mode network (DMN) decomposed into precuneus/posterior DMN and posterior DMN. The remaining iCAPs comprised anterior insula/amygdala and cerebellar network decomposed into anterior cerebellum/vermis, posterior cerebellum and cerebellum/visual networks (Figure 1 and Supplementary Table S1).

**Figure 1.**
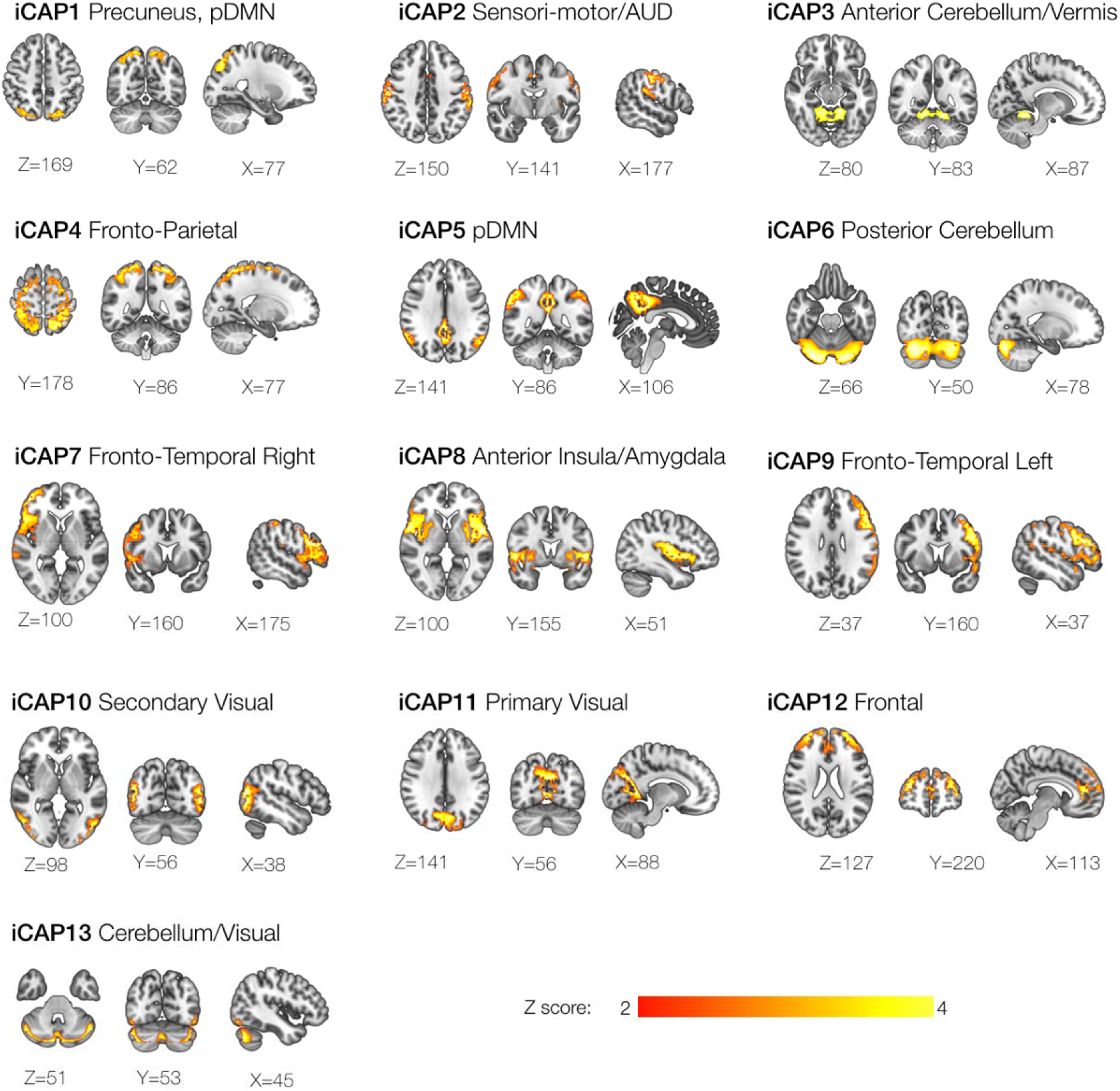
Spatial patterns of the 13 innovation-driven coactivation patterns (iCAPs) retrieved from all participants include: precuneus/posterior default mode network (DMN), sensori-motor/auditory (AUD), anterior cerebellum/vermis, fronto-parietal, posterior default mode network (pDMN), posterior cerebellum, fronto-temporal right, anterior insula/amygdala, fronto-temporal left, secondary visual, primary visual, frontal, cerebellum/visual. Locations denote displayed slices in Montreal Neurological Institute coordinates.

**Figure 1.**
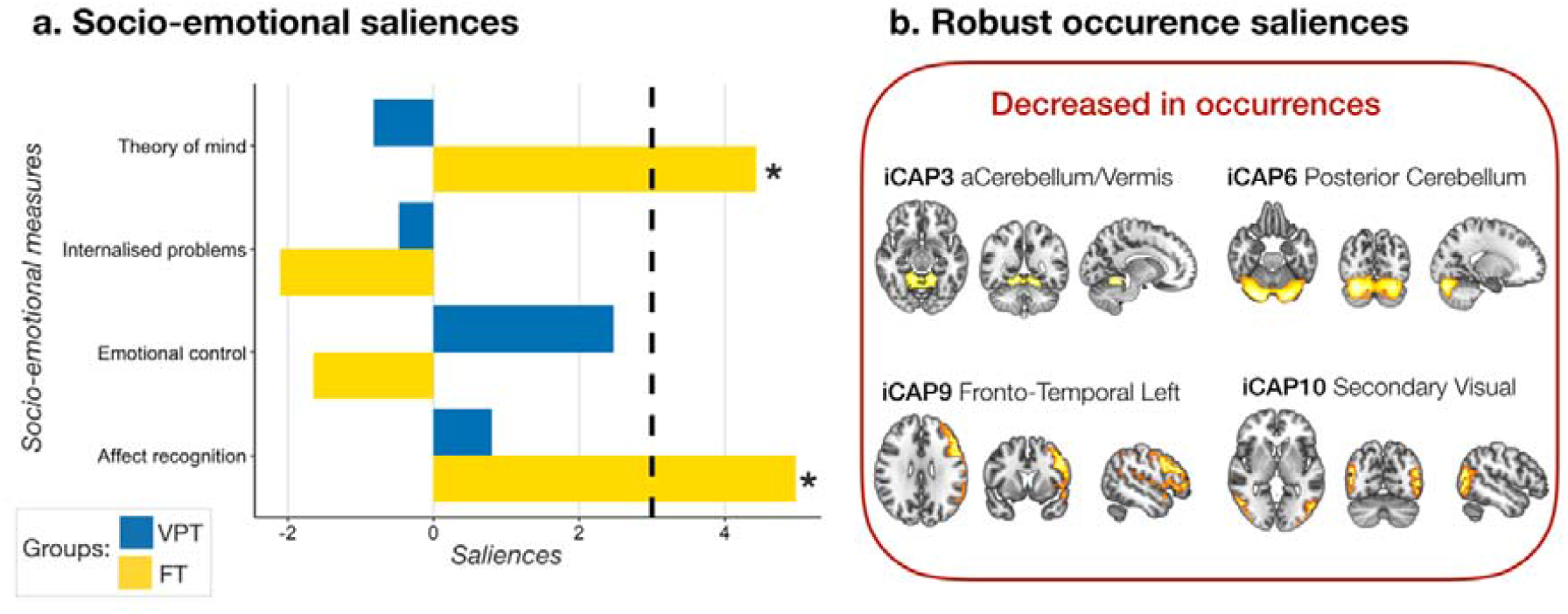
Associations between socio-emotional abilities and occurrence of the 13 iCAPs in the very preterm (VPT) and full-term (FT) groups based on the PLSC analysis. The diverging graph shows bootstrap ratio z-scores (x-axis) for each socio-emotional measures (y-axis) for the VPT and the FT groups in blue and yellow, respectively. Socio-emotional measures with an absolute bootstrap ratio z-score ≥ 3 or ≤ −3 (i.e., indicated by a back dash-dotted line on the graph) yield a robust contribution to the component (i.e., indicated by a black star). Only robust bootstrap ratio z-scores (absolute bootstrap ratio z-scores above or equal to 3) for iCAPs’ occurrences are shown.

### Group comparison of temporal properties of networks

We compared the VPT and the full-term groups for each temporal property considered, i.e., the occurrence and the total duration for each iCAPs, as well as the percent joint activation time of coupling and anticoupling for each pair of iCAPs. There was no significant group difference for any of the temporal measures after multiple comparison correction (Supplementary Tables S2, S3, S4, S5).

### Group comparison of laterality of networks

We compared the VPT and the full-term groups for the A-LI measure. There was no significant group difference for the laterality measure after multiple comparison correction (Supplementary Table S6).

### Association between iCAPs’ dynamics and socio-emotional measures

First, the PLSC analysis applied on socio-emotional scores and occurrence of the 13 iCAPs in the preterm and the full-term controls groups identified one statistically significant latent components: latent component 1 (p = 0.006). Latent component 1 revealed a pattern of significant association in the full-term group only (Figure 1). In full-term children, increased socio-emotional abilities (i.e., affect recognition and theory of mind) were associated with increased occurrence of iCAP3-anterior cerebellum/vermis, iCAP6-posterior cerebellum, iCAP9-fronto-temporal left, iCAP10-visual secondary. In the VPT group, there was no significant association between socio-emotional abilities and occurrences of the 13 iCAPs network.

Second, the PLSC analysis applied on socio-emotional measures and the total duration of the 13 iCAPs in the VPT and the full-term groups show no significant latent component.

Third, the PLSC analysis applied on socio-emotional measures and coupling duration of the 13 iCAPs in the VPT and the full-term groups identified one statistically significant latent components: latent component 1 (p = 0.002). Latent component 1 revealed a pattern of significant association in the VPT group only (Figure 2). In VPT children, increased socio-emotional abilities (i.e., decreased difficulties in emotional control and internalised problems) were associated with patterns of increase and decrease duration of coupling between the different iCAP networks. More specifically, in VPT, better socio-emotional abilities were associated with increased coupling duration between iCAPS 8 to 12 (anterior insula/amygdala to frontal network), iCAPS 9 to 10 (fronto-temporal left to secondary visual), iCAPS 4 to 6 (fronto-parietal to posterior cerebellum) and iCAPS 1 to 3 (precuneus, pDMN to anterior cerebellum/vermis); and with decreased coupling duration between iCAPS 5 to 8 (posterior DMN to anterior insula/amygdala), iCAPS 1 to 6 (precuneus, pDMN to posterior cerebellum), iCAPS 6 to 9 (posterior cerebellum to fronto-temporal left), iCAPS 2 to 9 (sensori-motor/auditory to fronto-temporal left), iCAPS 9 to 13 (fronto-temporal left to cerebellum/visual), iCAPS 11 to 13 (primary visual to cerebellum/visual), iCAPS 5 to 10 (posterior DMN to secondary visual), iCAPS 4 to 10 (fronto-parietal to secondary visual), iCAPS 6 to 13 (posterior cerebellum to cerebellum/visual), iCAPS 9 to 12 (fronto-temporal left to frontal), iCAPS 2 to 13 (sensori-motor/auditory to cerebellum/visual), iCAPS 2 to 3 (sensori-motor/auditory to 15 anterior cerebellum/vermis), iCAPS 8 to 10 (anterior insula/amygdala to secondary visual), iCAPS 2 to 12 (sensori-motor/auditory to frontal).

**Figure 2.**
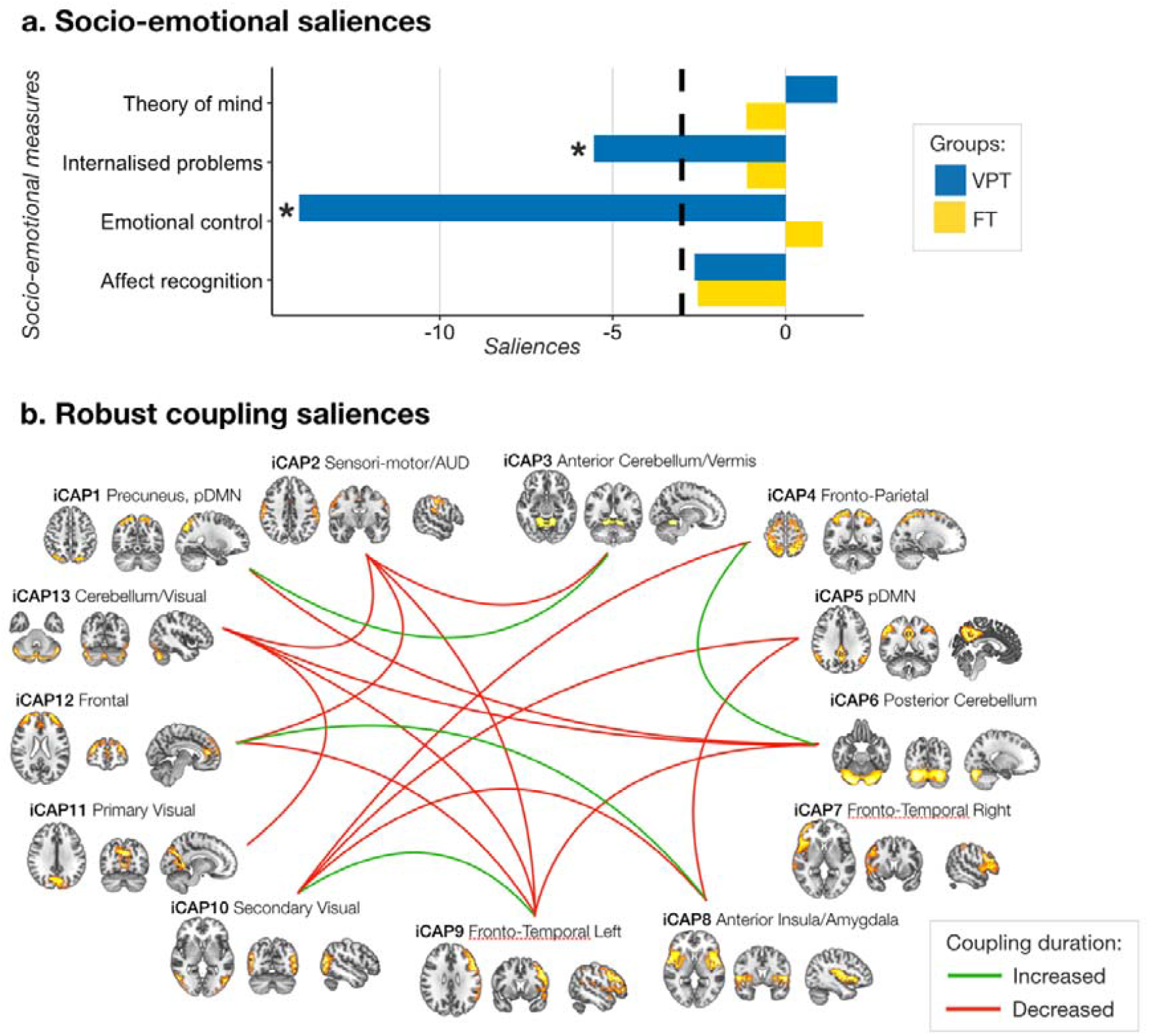
Associations between socio-emotional abilities and coupling of the 13 pairs of iCAPs in the very preterm (VPT) and full-term (FT) groups based on the PLSC analysis. The diverging graph shows bootstrap ratio z-scores (x-axis) for each socio-emotion measure (y-axis) for the VPT and the FT groups in blue and yellow, respectively. Socio-emotional measures with an absolute bootstrap ratio z-score ≥ 3 or ≤ −3 (i.e., indicated by a back dash-dotted line on the graph) yield a robust contribution to the component (i.e., indicated by a black star). Only robust bootstrap ratio z-scores (absolute bootstrap ratio z-scores ≥ 3 or ≤ −3) for iCAPs’ couplings are shown.

Finally, the PLSC analysis applied on socio-emotional measures and anticoupling duration of the 13 iCAPs in the VPT and the full-term groups show no significant latent component.

Original saliences as well as their bootstrap-estimated standard deviations and bootstrap ratio z-scores for the PLSC analyses showing a significant latent component are reported in Supplementary Table S8 and S9.

## Discussion

Using a novel whole-brain data-driven approach tailored to disentangle transient activity of coordinated brain networks from spontaneous rs-fMRI measurements, we unravelled large-scale resting state dynamics in VPT and full-term children aged 6 to 9 years. In this cohort of well-functioning VPT children, spatial organisation of the 13 networks retrieved was comparable to full-term controls. Dynamic features and lateralisation of network brain activity were also comparable across groups for the 13 large-scale brain networks. Despite apparent similarities in terms of dynamical FC parameters, multivariate pattern analysis unveiled group differences in their associations with socio-emotional abilities. While a pattern of decreased engagement in certain brain networks were associated with better socio-emotional abilities in full-term controls; in the VPT group, better socio-emotional abilities were associated with coordination of activity across different networks, i.e., coupling duration between different pairs of networks.

In this cohort of well-functioning VPT children, overall dynamic FC characteristics were comparable to the full-term control group. First, spatial organisation of spontaneous brain activity was similar across the two groups. Second, temporal dynamics of iCAPs brain network were comparable in terms of occurrence and duration of engagement in a given network as well as in terms of coupling and anticoupling between pairs of networks. Moreover, the lateralisation of the large-scale brain networks retrieved was also comparable across groups. This pattern of dynamic FC similarities between VPT and full-term children are not consistent with previous studies using static FC (Degnan et al., 2015; Mossad et al., 2021; Rowlands et al., 2016; Wehrle et al., 2018). To the best of our knowledge, there is currently no study in school-age preterm children exploring dynamic FC. Nevertheless, Stoecklein and colleagues (2020) recently used FC variability or moment-to-moment variations of BOLD signals that represent neural dynamic range or flexibility completed in preterm and full-term infants (Stoecklein et al., 2020). Whole-brain FC variability was highly similar between 50 preterm and 25 full-term infants at term equivalent age suggesting that prematurity does not influence significantly FC variability at term equivalent age.

In the current study, children with an IQ below 70 as well as children with high rates of motion artefact in rs-fMRI time series were excluded. It is possible that only well-functioning children with good self-regulation abilities were included in the final analyses. A recent study from Bolton and colleagues (2020) specifically examined motion characteristics during rs-fMRI acquisition in healthy adults (Bolton et al., 2020). Results show a broad array of behavioural and clinical characteristics related to motion including, among other, self-regulation, anxiety and depression. During childhood, it is possible that the association between motion and behavioural characteristics might be even more exacerbate resulting in the exclusion of children with increased behavioural difficulties in daily life and therefore increased atypicality in functional brain organisation.

The iCAPs framework has previously being found to provide relevant makers of psychiatric symptomatology in adults and children, such as depression, anxiety and psychotic symptoms (Piguet et al., 2021; Zöller et al., 2019). In the current study, despite comparable dynamic features of resting state networks between VPT and full-term controls, different patterns of association with socio-emotional outcomes were observed across the two groups. In full-term controls, increased socio-emotional abilities were associated with reduced occurrence of different resting state networks, including the anterior cerebellum/vermis, posterior cerebellum, fronto-temporal left, and visual secondary networks. At the opposite, in VPT children, increased socio-emotional abilities were associated with a large pattern of increased and decreased coupling between networks. This is in line with previous studies conducted in the VPT population, suggesting that cognitive abilities rely more on coupling between networks in comparison to full-term controls for whom cognitive abilities is more related to the involvement of a given network (Barnes-Davis et al., 2018; Bäuml et al., 2015; Degnan et al., 2015; Mossad et al., 2021). Altogether, the findings suggest that, while in the full-term control group socio-emotional abilities was associated with different degree of integration of functional brain networks, i.e., recruitment of intra-network FC; in the VPsT group, socio-emotional abilities were associated with different degree of segregation between networks, i.e. increase and decrease in recruitment of inter-network FC. According to previous studies, typical development of functional architecture appears to show ongoing changes with an increase in integration along with a decrease in segregation of functional networks as children get older (Dosenbach et al., 2010; Menon, 2013; Rohr et al., 2018). Structurally this increased segregation and loss of increased integration has been shown at different ages in preterm compare to full-term infants (de Almeida et al., 2021; Fischi-Gomez et al., 2016). It is therefore possible that the group difference observed in the pattern of association between dynamic FC features and socio-emotional abilities reflect reduced degree of maturation of functional architecture in the VPT group.

While providing new insight into dynamic functional organisaton in children born VPT, the current study has a number of limitations which need to be considered. First, the sample size of the study is rather small and warrants validation in larger samples. The small sample size may have limited the identification of subtle group differences in FC dynamics. Moreover, study participants comprised only well-functioning VPT children. The exclusion of 13 participants due to significant head motion during rs-fMRI acquisition might have accentuate this point even more through the exclusion of children with increased behavioural difficulties. Taking these limitations into account, dynamic features of FC might represent a relevant neuroimaging markers and inform on potential mechanisms through which preterm birth leads to neurodevelopmental deficits.

## Conclusions

Leveraging recent advances in analysis of dynamic features of FC, we explored precise moments of brain network activation and interaction in VPT compared to full-term children from 6 to 9 years of age. In the current study, brain networks were comparable across groups not only in terms of spatial organisation but also in terms of laterality and dynamic characteristics. Despite group similarities in dynamical FC parameters, multivariate pattern analysis revealed different pattern of association with socio-emotional abilities in the VPT and full-term groups, that might reflect reduced degree of maturation of functional architecture in the VPT group.

## Data Availability

Deidentified individual participant data (including data dictionaries) will be made available, in addition to study protocols, the statistical analysis plan, and the informed consent form. The data will be made available upon publication to researchers who provide a methodologically sound proposal for use in achieving the goals of the approved proposal. Proposals should be submitted to.

## Acknowledgements

We thank and acknowledge all participating young adolescents and families who made this research possible. We also thank the Fondation Campus Biotech Geneva (FCBG), a foundation of the Swiss Federal Institute of Technology Lausanne (EPFL), the University of Geneva (UniGe), and the University Hospitals of Geneva (HUG); the Research Platform of the University Hospitals of Geneva (HUG) for their practical help.

