## Supplementary Materials for "Large scale brain network dynamics in very preterm children and relationship with socio-emotional outcomes"

### Supplementary figures

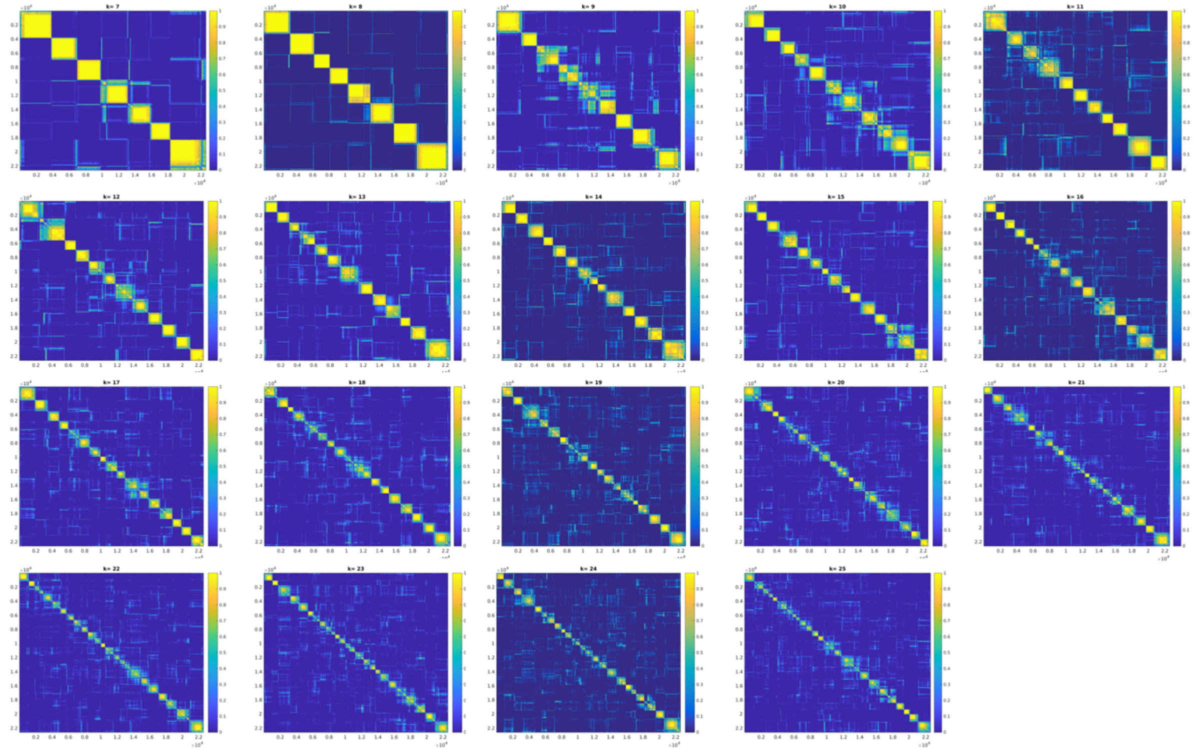

**Figure S1**. Consensus matrices for cluster numbers K = 7 to K = 25. High values in the matrix indicate that the two corresponding frames were clustered together during re-sampling. A value of 1 means that the frames were always clustered together, while a value of 0 means that the frames were never clustered together. We selected to use K = 13 based on visual inspection of the consensus matrices and evaluation of consensus clustering quality measures (Monti et al., 2003). In Supplementary Figure S2, we plot the distribution of average consensus values per cluster to assess the quality of the clustering for each K.

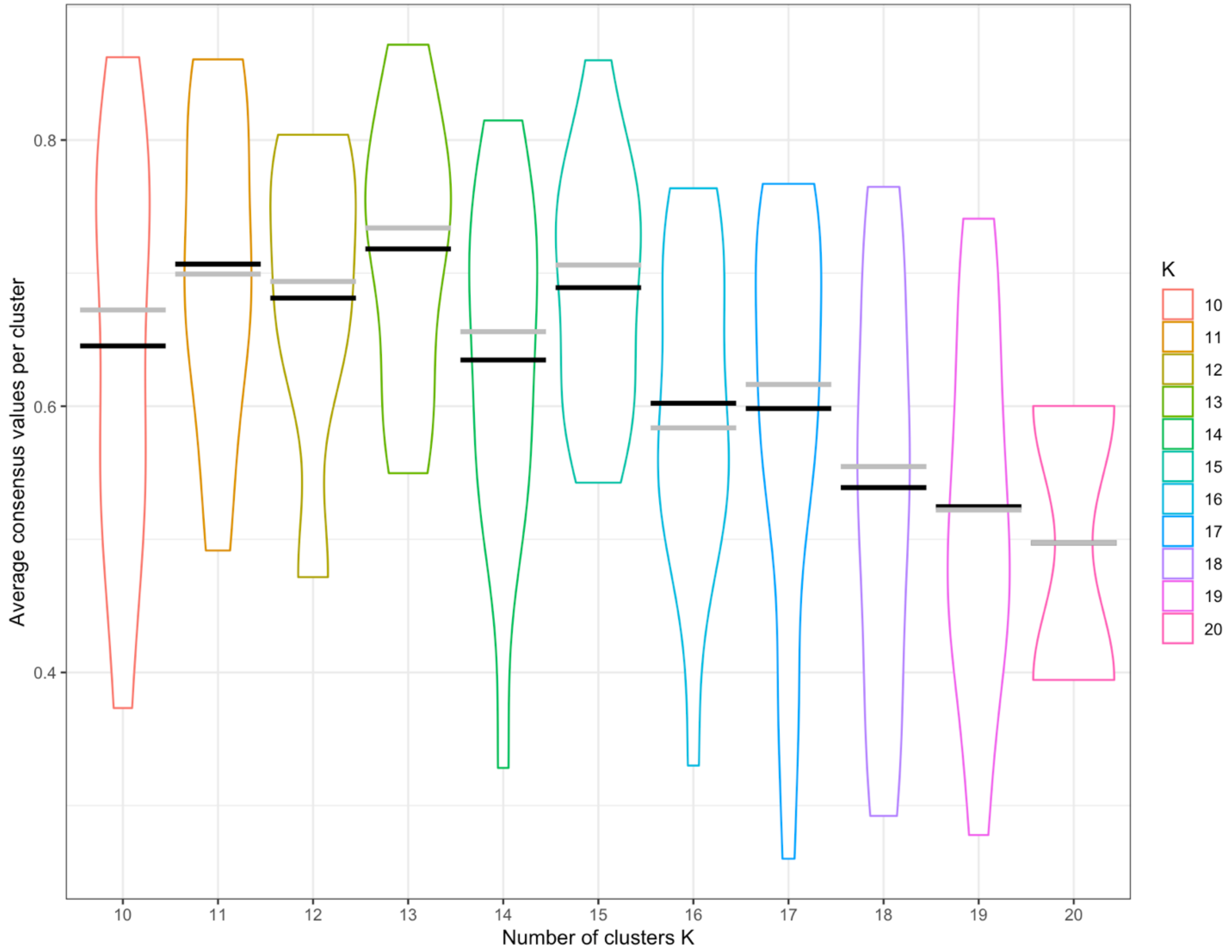


**Figure S2**. Consensus clustering quality measures for cluster numbers K = 10 to K = 20. The distribution of average consensus values per cluster: the mean for each cluster is represented by a black line and the median is represented by a grey line. Mean consensus is maximal (closest to 1) for K = 13.

### Supplementary Tables

**Table S1**. iCAPs functional networks of regions from the automated anatomical labeling 2 (AAL2) atlas. Percentiles indicate the fraction of voxels of a functional network or region that have a z-score > 2.3. A network/region is listed if more than 20% of the network/region is included in the iCAP.

| iCAP | Lobe | Region | Percentile | mean z-score | voxels |
|---|---|---|---|---|---|
| **iCAP 1** | Parietal | Parietal_Sup_R | 64.82 | 2.14 | 457 |
| | Parietal | Parietal_Sup_L | 64.6 | 2.12 | 480 |
| | Occipital | Occipital_Sup_L | 52.4 | 2.11 | 164 |
| | Occipital | Occipital_Sup_R | 50.49 | 2.18 | 154 |
| | Parietal | Angular_R | 29.57 | 1.97 | 225 |
| | Posterior Fossa | Cerebelum_6_R | 27.52 | 1.8 | 368 |
| | Parietal | Precuneus_L | 23.9 | 1.92 | 365 |
| | Parietal | Precuneus_R | 23.21 | 1.84 | 331 |
| | Posterior Fossa | Cerebelum_Crus1_L | 19.67 | 1.71 | 302 |
| | Posterior Fossa | Cerebelum_Crus1_R | 18.27 | 1.72 | 273 |
| | Posterior Fossa | Cerebelum_6_L | 17.7 | 1.65 | 229 |
| | Parietal | Parietal_Inf_L | 16.4 | 1.84 | 208 |
| | Occipital | Cuneus_R | 14.68 | 1.75 | 74 |
| | Occipital | Occipital_Mid_R | 13.65 | 1.94 | 107 |
| | Occipital | Occipital_Mid_L | 12.91 | 2.05 | 143 |
| | Occipital | Cuneus_L | 11.24 | 1.78 | 79 |
| | Parietal | Parietal_Inf_R | 10.16 | 1.65 | 78 |
| **iCAP 2** | Parietal | SupraMarginal_L | 79.53 | 2.73 | 470 |
| | Central | Rolandic_Oper_L | 62.3 | 2.07 | 304 |
| | Parietal | SupraMarginal_R | 62.06 | 2.68 | 535 |
| | Frontal | Rolandic_Oper_R | 58.16 | 2.04 | 342 |
| | Central | Postcentral_L | 57.39 | 2.2 | 652 |

| | | | | | |
|---|---|---|---|---|---|
| | Central | Precentral_R | 57.39 | 1.96 | 621 |
| | Central | Postcentral_R | 49.44 | 2.48 | 613 |
| | Temporal | Heschl_L | 35.81 | 1.74 | 53 |
| | Central | Precentral_L | 35.13 | 1.82 | 372 |
| | Frontal | Supp_Motor_Area_R | 33.33 | 1.82 | 121 |
| | Temporal | Temporal_Sup_L | 30.44 | 2.12 | 309 |
| | Frontal | Supp_Motor_Area_L | 29.17 | 1.82 | 119 |
| | Limbic | Insula_L | 26.58 | 1.79 | 248 |
| | Temporal | Heschl_R | 25.76 | 1.68 | 17 |
| | Limbic | Insula_R | 24.33 | 1.75 | 236 |
| | Limbic | Cingulate_Mid_L | 21.08 | 1.81 | 219 |
| | Limbic | Cingulate_Mid_R | 20.57 | 1.81 | 224 |
| | Parietal | Parietal_Inf_L | 20.43 | 2.31 | 259 |
| | Frontal | Frontal_Inf_Oper_L | 16.27 | 1.75 | 61 |
| | Frontal | Frontal_Inf_Oper_R | 15.52 | 1.64 | 88 |
| | Temporal | Temporal_Sup_R | 10.06 | 1.87 | 104 |
| **iCAP 3** | Posterior Fossa | Cerebelum_4_5_R | 83.05 | 2.1 | 544 |
| | Posterior Fossa | Vermis_9 | 81.72 | 1.83 | 76 |
| | Posterior Fossa | Vermis_3 | 80.95 | 1.87 | 17 |
| | Posterior Fossa | Cerebelum_4_5_L | 75.31 | 2.05 | 540 |
| | Posterior Fossa | Vermis_4_5 | 67.33 | 2.01 | 204 |
| | Posterior Fossa | Cerebelum_9_R | 62.07 | 1.74 | 36 |
| | Posterior Fossa | Cerebelum_3_R | 54.84 | 1.97 | 17 |
| | Posterior Fossa | Vermis_8 | 46.58 | 1.63 | 68 |
| | Posterior Fossa | Cerebelum_3_L | 38.46 | 1.85 | 5 |
| | Posterior Fossa | Cerebelum_9_L | 36.36 | 1.67 | 20 |
| | Posterior Fossa | Vermis_7 | 27.01 | 1.6 | 37 |
| | Occipital | Fusiform_R | 22.68 | 1.83 | 276 |

| | | | | | |
|---|---|---|---|---|---|
| | Posterior Fossa | Vermis_6 | 20.97 | 1.65 | 56 |
| | Posterior Fossa | Cerebelum_6_L | 19.09 | 1.74 | 247 |
| | Posterior Fossa | Cerebelum_6_R | 15.56 | 1.74 | 208 |
| | Occipital | Fusiform_L | 14.74 | 1.82 | 175 |
| | Occipital | Lingual_R | 14.69 | 1.82 | 144 |
| | Posterior Fossa | Vermis_1_2 | 12.5 | 1.71 | 1 |
| | Occipital | Lingual_L | 10.48 | 1.79 | 103 |
| **iCAP 4** | Parietal | Parietal_Sup_L | 99.06 | 2.51 | 736 |
| | Parietal | Parietal_Sup_R | 99.01 | 2.35 | 698 |
| | Frontal | Paracentral_Lobule_R | 92.68 | 2.02 | 76 |
| | Frontal | Paracentral_Lobule_L | 88.69 | 2.41 | 149 |
| | Parietal | Parietal_Inf_R | 84.11 | 2.03 | 646 |
| | Frontal | Supp_Motor_Area_R | 83.2 | 2.1 | 302 |
| | Parietal | Parietal_Inf_L | 80.84 | 2.02 | 1025 |
| | Central | Postcentral_R | 70.73 | 2.32 | 877 |
| | Frontal | Supp_Motor_Area_L | 69.85 | 2.11 | 285 |
| | Central | Postcentral_L | 51.41 | 2.4 | 584 |
| | Central | Precentral_R | 48.24 | 2.19 | 522 |
| | Occipital | Occipital_Sup_R | 43.28 | 1.87 | 132 |
| | Parietal | Angular_R | 43.23 | 1.93 | 329 |
| | Central | Precentral_L | 40.13 | 2.22 | 425 |
| | Occipital | Occipital_Sup_L | 39.3 | 1.89 | 123 |
| | Parietal | Precuneus_L | 37 | 2.31 | 565 |
| | Frontal | Frontal_Sup_2_R | 34.5 | 2.41 | 376 |
| | Parietal | Precuneus_R | 30.86 | 1.98 | 440 |
| | Frontal | Frontal_Sup_2_L | 24.28 | 2.3 | 219 |
| | Parietal | Angular_L | 21.94 | 1.8 | 129 |
| | Limbic | Cingulate_Mid_R | 20.48 | 1.63 | 223 |

| | | | | | |
|---|---|---|---|---|---|
| | Parietal | SupraMarginal_L | 14.72 | 1.73 | 87 |
| | Limbic | Cingulate_Mid_L | 14.53 | 1.59 | 151 |
| | Occipital | Occipital_Mid_L | 12.18 | 1.86 | 135 |
| | Parietal | SupraMarginal_R | 10.9 | 1.75 | 94 |
| | Occipital | Cuneus_R | 10.71 | 1.69 | 54 |
| **iCAP 5** | Frontal | Frontal_Med_Orb_L | 100 | 1.53 | 1 |
| | Limbic | Cingulate_Post_L | 96.28 | 3.15 | 181 |
| | Parietal | Angular_L | 92.52 | 2.96 | 544 |
| | Limbic | Cingulate_Post_R | 89.16 | 3.07 | 74 |
| | Parietal | Angular_R | 79.89 | 2.6 | 608 |
| | Parietal | Parietal_Inf_R | 70.96 | 2.72 | 545 |
| | Parietal | Parietal_Inf_L | 62.22 | 2.72 | 789 |
| | Parietal | Precuneus_L | 54.94 | 2.64 | 839 |
| | Parietal | Precuneus_R | 45.93 | 2.69 | 655 |
| | Limbic | Cingulate_Mid_L | 43.89 | 2.59 | 456 |
| | Limbic | Cingulate_Mid_R | 41.41 | 2.72 | 451 |
| | Frontal | Frontal_Med_Orb_R | 33.33 | 1.6 | 15 |
| | Parietal | SupraMarginal_R | 27.96 | 2.22 | 241 |
| | Parietal | Parietal_Sup_L | 19.92 | 2.22 | 148 |
| | Temporal | Temporal_Mid_L | 17.62 | 1.7 | 399 |
| | Parietal | Parietal_Sup_R | 15.32 | 2.09 | 108 |
| | Parietal | SupraMarginal_L | 10.49 | 1.91 | 62 |
| | Limbic | Cingulate_Ant_L | 10.08 | 1.6 | 79 |
| **iCAP 6** | Posterior Fossa | Cerebelum_7b_R | 100 | 2.17 | 46 |
| | Posterior Fossa | Cerebelum_8_R | 98.75 | 1.99 | 79 |
| | Posterior Fossa | Cerebelum_Crus1_R | 97.52 | 2.65 | 1457 |
| | Posterior Fossa | Cerebelum_Crus1_L | 95.77 | 2.51 | 1470 |
| | Posterior Fossa | Vermis_8 | 95.21 | 1.93 | 139 |

| | | | | | |
|---|---|---|---|---|---|
| | Posterior Fossa | Cerebelum_6_R | 91.4 | 2.42 | 1222 |
| | Posterior Fossa | Cerebelum_9_L | 90.91 | 1.77 | 50 |
| | Posterior Fossa | Cerebelum_7b_L | 90.22 | 1.9 | 83 |
| | Posterior Fossa | Cerebelum_Crus2_L | 89.57 | 2.21 | 713 |
| | Posterior Fossa | Cerebelum_Crus2_R | 88.65 | 2.29 | 609 |
| | Posterior Fossa | Vermis_7 | 88.32 | 2.21 | 121 |
| | Posterior Fossa | Vermis_9 | 88.17 | 1.91 | 82 |
| | Posterior Fossa | Cerebelum_6_L | 82.3 | 2.23 | 1065 |
| | Posterior Fossa | Cerebelum_8_L | 81.25 | 1.75 | 52 |
| | Posterior Fossa | Cerebelum_9_R | 74.14 | 1.81 | 43 |
| | Posterior Fossa | Vermis_6 | 60.67 | 1.91 | 162 |
| | Occipital | Occipital_Inf_R | 53.15 | 1.81 | 118 |
| | Occipital | Fusiform_R | 40.51 | 2.13 | 493 |
| | Occipital | Lingual_R | 36.84 | 1.97 | 361 |
| | Occipital | Fusiform_L | 33.36 | 2 | 396 |
| | Occipital | Lingual_L | 20.85 | 1.88 | 205 |
| | Temporal | Temporal_Inf_R | 13.32 | 1.78 | 151 |
| | Occipital | Occipital_Inf_L | 12.5 | 1.64 | 31 |
| | Temporal | Temporal_Inf_L | 10.61 | 1.75 | 89 |
| **iCAP 7** | Frontal | Frontal_Inf_Tri_R | 99.7 | 3.12 | 655 |
| | Frontal | Frontal_Inf_Oper_R | 99.12 | 2.95 | 562 |
| | Central | Frontal_Inf_Orb_2_R | 92.93 | 2.25 | 92 |
| | Frontal | Frontal_Mid_2_R | 91.08 | 2.78 | 1593 |
| | Frontal | Rolandic_Oper_R | 81.12 | 2.02 | 477 |
| | Temporal | Temporal_Pole_Mid_R | 78.57 | 1.82 | 22 |
| | Parietal | SupraMarginal_R | 74.83 | 1.86 | 645 |
| | Limbic | Insula_R | 73.09 | 2.1 | 709 |
| | Temporal | Temporal_Sup_R | 72.05 | 1.92 | 745 |

| | | | | | |
|---|---|---|---|---|---|
| | Temporal | Temporal_Pole_Sup_R | 69.19 | 1.97 | 128 |
| | Frontal | OFCpost_R | 66.67 | 1.55 | 4 |
| | Frontal | Frontal_Sup_2_R | 56.97 | 2.22 | 621 |
| | Subcortical grey nucleus | Putamen_R | 45.15 | 1.77 | 279 |
| | Temporal | Heschl_R | 43.94 | 1.98 | 29 |
| | Parietal | Parietal_Inf_R | 39.58 | 1.93 | 304 |
| | Central | Postcentral_R | 38.15 | 1.87 | 473 |
| | Temporal | Temporal_Mid_R | 36.18 | 1.82 | 683 |
| | Central | Precentral_R | 35.95 | 2.2 | 389 |
| | Frontal | Frontal_Sup_Medial_R | 20.39 | 1.65 | 73 |
| | Temporal | Temporal_Inf_R | 11.2 | 1.61 | 127 |
| | Central | Rolandic_Oper_L | 98.57 | 2.8 | 481 |
| | Limbic | Insula_R | 98.56 | 2.55 | 956 |
| | Limbic | Insula_L | 98.07 | 2.66 | 915 |
| | Temporal | Heschl_L | 97.97 | 2.58 | 145 |
| **iCAP 8** | Frontal | Rolandic_Oper_R | 97.79 | 2.58 | 575 |
| | Temporal | Heschl_R | 96.97 | 2.3 | 64 |
| | Temporal | Temporal_Pole_Sup_R | 92.97 | 2.35 | 172 |
| | Temporal | Temporal_Pole_Mid_R | 92.86 | 2.03 | 26 |
| | Temporal | Temporal_Pole_Sup_L | 90.07 | 2.28 | 127 |
| | Subcortical grey nucleus | Putamen_R | 86.89 | 2.17 | 537 |
| | Subcortical grey nucleus | Putamen_L | 83.92 | 2.21 | 407 |
| | Frontal | OFCpost_R | 83.33 | 1.83 | 5 |
| | Subcortical grey nucleus | Amygdala_R | 82.41 | 1.95 | 89 |
| | Temporal | Temporal_Sup_L | 80.99 | 2.23 | 822 |
| | Frontal | Frontal_Inf_Orb_2_L | 80.67 | 1.99 | 121 |
| | Central | Frontal_Inf_Orb_2_R | 77.78 | 1.94 | 77 |
| | Subcortical grey nucleus | Amygdala_L | 62.6 | 1.86 | 77 |

| | | | | | |
|---|---|---|---|---|---|
| | Frontal | OFCpost_L | 62.5 | 1.69 | 5 |
| | Limbic | Hippocampus_R | 53.23 | 1.77 | 107 |
| | Subcortical grey nucleus | Pallidum_R | 51.85 | 1.87 | 28 |
| | Temporal | Temporal_Sup_R | 50.39 | 2.02 | 521 |
| | Frontal | Frontal_Inf_Tri_R | 46.58 | 1.99 | 306 |
| | Frontal | Frontal_Inf_Oper_L | 44.53 | 2.05 | 167 |
| | Frontal | Frontal_Inf_Tri_L | 38.62 | 2.09 | 297 |
| | Frontal | Frontal_Inf_Oper_R | 34.04 | 2.26 | 193 |
| | Frontal | Olfactory_R | 28.67 | 1.91 | 43 |
| | Limbic | Hippocampus_L | 28.02 | 1.7 | 72 |
| | Limbic | ParaHippocampal_R | 25.97 | 1.78 | 120 |
| | Parietal | SupraMarginal_L | 23.01 | 2.16 | 136 |
| | Subcortical grey nucleus | Pallidum_L | 21.74 | 1.78 | 10 |
| | Parietal | SupraMarginal_R | 17.52 | 2.03 | 151 |
| | Frontal | Olfactory_L | 16.33 | 1.88 | 16 |
| | Subcortical grey nucleus | Caudate_R | 13.39 | 1.61 | 32 |
| | Central | Postcentral_L | 10.56 | 2.22 | 120 |
| **iCAP 9** | Frontal | Frontal_Inf_Tri_L | 99.22 | 2.76 | 763 |
| | Frontal | Frontal_Inf_Oper_L | 98.93 | 2.67 | 371 |
| | Frontal | Frontal_Mid_2_L | 95.41 | 2.33 | 1434 |
| | Frontal | Frontal_Inf_Orb_2_L | 95.33 | 2.16 | 143 |
| | Central | Rolandic_Oper_L | 91.8 | 1.92 | 448 |
| | Parietal | SupraMarginal_L | 88.16 | 1.99 | 521 |
| | Temporal | Temporal_Sup_L | 87.68 | 2.13 | 890 |
| | Temporal | Temporal_Pole_Sup_L | 79.43 | 2.16 | 112 |
| | Limbic | Insula_L | 78.03 | 1.91 | 728 |
| | Temporal | Heschl_L | 75.68 | 1.84 | 112 |
| | Frontal | OFCpost_L | 75 | 1.69 | 6 |

| | | | | | |
|---|---|---|---|---|---|
| | Temporal | Temporal_Mid_L | 69.52 | 2.15 | 1574 |
| | Central | Precentral_L | 66.1 | 2.51 | 700 |
| | Frontal | Frontal_Sup_2_L | 64.19 | 1.89 | 579 |
| | Parietal | Parietal_Inf_L | 53.71 | 1.84 | 681 |
| | Central | Postcentral_L | 52.64 | 1.93 | 598 |
| | Parietal | Angular_L | 47.45 | 1.83 | 279 |
| | Subcortical grey nucleus | Putamen_L | 47.01 | 1.71 | 228 |
| | Frontal | Frontal_Sup_Medial_L | 22.36 | 1.75 | 108 |
| | Temporal | Temporal_Inf_L | 21.1 | 1.68 | 177 |
| **iCAP 10** | Occipital | Occipital_Mid_R | 88.39 | 3.1 | 693 |
| | Occipital | Occipital_Mid_L | 82.58 | 3.1 | 915 |
| | Occipital | Occipital_Inf_L | 71.77 | 2.34 | 178 |
| | Temporal | Temporal_Mid_R | 55.56 | 2.79 | 1049 |
| | Occipital | Occipital_Inf_R | 49.55 | 2.02 | 110 |
| | Occipital | Occipital_Sup_L | 43.77 | 2.35 | 137 |
| | Temporal | Temporal_Mid_L | 41.03 | 2.31 | 929 |
| | Occipital | Occipital_Sup_R | 31.15 | 2.26 | 95 |
| | Occipital | Cuneus_L | 27.45 | 1.79 | 193 |
| | Temporal | Temporal_Sup_R | 27.27 | 2.05 | 282 |
| | Parietal | Angular_R | 26.68 | 1.97 | 203 |
| | Parietal | Angular_L | 21.77 | 1.89 | 128 |
| | Temporal | Temporal_Inf_R | 21.16 | 2.09 | 240 |
| | Occipital | Cuneus_R | 18.45 | 1.74 | 93 |
| | Occipital | Calcarine_L | 11.05 | 1.69 | 121 |
| | Temporal | Temporal_Sup_L | 11.03 | 1.85 | 112 |
| | Temporal | Temporal_Inf_L | 10.25 | 1.93 | 86 |
| | Posterior Fossa | Cerebelum_Crus2_R | 10.19 | 1.64 | 70 |
| | Occipital | Calcarine_R | 10.06 | 1.57 | 63 |

| | | | | | |
|---|---|---|---|---|---|
| **iCAP 11** | Occipital | Cuneus_L | 99.86 | 2.98 | 702 |
| | Occipital | Cuneus_R | 94.84 | 2.61 | 478 |
| | Occipital | Calcarine_R | 94.09 | 3.62 | 589 |
| | Occipital | Occipital_Sup_L | 84.66 | 2.06 | 265 |
| | Occipital | Calcarine_L | 81.28 | 3.46 | 890 |
| | Occipital | Lingual_L | 69.68 | 2.48 | 685 |
| | Occipital | Lingual_R | 55.2 | 2.33 | 541 |
| | Occipital | Occipital_Sup_R | 55.08 | 1.82 | 168 |
| | Occipital | Occipital_Mid_R | 43.49 | 1.76 | 341 |
| | Parietal | Precuneus_R | 43.27 | 2.79 | 617 |
| | Occipital | Occipital_Mid_L | 42.96 | 1.88 | 476 |
| | Parietal | Precuneus_L | 31.83 | 2.55 | 486 |
| | Posterior Fossa | Cerebelum_4_5_L | 11.44 | 1.92 | 82 |
| **iCAP 12** | Frontal | Frontal_Med_Orb_L | 100 | 4.19 | 1 |
| | Frontal | Frontal_Med_Orb_R | 95.56 | 3.19 | 43 |
| | Limbic | Cingulate_Ant_R | 94.95 | 3.05 | 470 |
| | Frontal | Frontal_Sup_Medial_R | 88.27 | 3.05 | 316 |
| | Limbic | Cingulate_Ant_L | 86.73 | 3.07 | 680 |
| | Frontal | Frontal_Sup_Medial_L | 79.5 | 2.6 | 384 |
| | Frontal | Frontal_Inf_Orb_2_L | 75.33 | 1.84 | 113 |
| | Frontal | Frontal_Sup_2_L | 65.96 | 3.04 | 595 |
| | Frontal | Frontal_Inf_Tri_L | 65.15 | 2.21 | 501 |
| | Frontal | Frontal_Mid_2_L | 63.87 | 2.68 | 960 |
| | Frontal | Frontal_Mid_2_R | 61.58 | 2.64 | 1077 |
| | Frontal | Frontal_Sup_2_R | 54.5 | 3.34 | 594 |
| | Frontal | Frontal_Inf_Tri_R | 49.01 | 2.1 | 322 |
| | Central | Frontal_Inf_Orb_2_R | 40.4 | 2.22 | 40 |
| **iCAP 13** | Posterior Fossa | Cerebelum_Crus2_R | 87.19 | 3.47 | 599 |

| | | | | |
|---|---|---|---|---|
| Posterior Fossa | Cerebelum_Crus2_L | 86.06 | 3.58 | 685 |
| Posterior Fossa | Vermis_8 | 84.93 | 2.74 | 124 |
| Occipital | Occipital_Inf_R | 79.28 | 2.27 | 176 |
| Posterior Fossa | Cerebelum_8_L | 67.19 | 2.09 | 43 |
| Posterior Fossa | Cerebelum_8_R | 66.25 | 2.03 | 53 |
| Occipital | Occipital_Inf_L | 63.31 | 2.25 | 157 |
| Posterior Fossa | Cerebelum_7b_R | 63.04 | 2.22 | 29 |
| Posterior Fossa | Vermis_7 | 53.28 | 2.02 | 73 |
| Posterior Fossa | Cerebelum_7b_L | 53.26 | 2.77 | 49 |
| Posterior Fossa | Cerebelum_Crus1_R | 52.28 | 2.94 | 781 |
| Posterior Fossa | Cerebelum_Crus1_L | 51.99 | 2.83 | 798 |
| Posterior Fossa | Vermis_9 | 38.71 | 2.17 | 36 |
| Temporal | Temporal_Inf_R | 27.95 | 1.85 | 317 |
| Posterior Fossa | Cerebelum_9_L | 27.27 | 1.86 | 15 |
| Posterior Fossa | Cerebelum_9_R | 24.14 | 2.09 | 14 |
| Occipital | Lingual_R | 20.2 | 2.57 | 198 |
| Posterior Fossa | Vermis_6 | 18.73 | 1.99 | 50 |
| Occipital | Calcarine_L | 17.72 | 2.14 | 194 |
| Temporal | Temporal_Inf_L | 17.16 | 1.8 | 144 |
| Occipital | Occipital_Mid_L | 13.9 | 1.89 | 154 |
| Occipital | Fusiform_L | 12.97 | 2.06 | 154 |
| Occipital | Lingual_L | 12.11 | 2.71 | 119 |

**Table S2**. Group comparison for the VPT and full-term control groups for the occurrence of the identified iCAPs networks

| | **VPT group** | | **Full-term group** | | | | **Group comparison** | | |
|---|---|---|---|---|---|---|---|---|---|
| **iCAPs – Occurrence** | **Mean** | **SD** | **Mean_FT** | **SD_FT** | **t** | **df** | **p-value** | **q-value (fdr)** | **Effect size (d)** |
| iCAP 1 | 34.679 | 5.863 | 34.333 | 3.774 | 0.222 | 31.705 | 0.826 | 0.948 | 0.065 |
| iCAP 2 | 28.179 | 6.301 | 28.583 | 4.814 | -0.221 | 27.127 | 0.827 | 0.948 | -0.068 |
| iCAP 3 | 24.179 | 6.037 | 24.583 | 7.879 | -0.159 | 16.799 | 0.876 | 0.948 | -0.061 |
| iCAP 4 | 17.286 | 3.670 | 17.333 | 4.559 | -0.032 | 17.411 | 0.975 | 0.975 | -0.012 |
| iCAP 5 | 20.357 | 6.442 | 24.083 | 6.288 | -1.705 | 21.361 | 0.103 | 0.948 | -0.582 |
| iCAP 6 | 23.321 | 6.481 | 24.750 | 4.159 | -0.833 | 31.786 | 0.411 | 0.948 | -0.242 |
| iCAP 7 | 17.393 | 4.856 | 17.000 | 5.908 | 0.203 | 17.687 | 0.842 | 0.948 | 0.076 |
| iCAP 8 | 14.893 | 5.370 | 13.333 | 5.483 | 0.829 | 20.494 | 0.416 | 0.948 | 0.289 |
| iCAP 9 | 20.214 | 5.776 | 21.667 | 5.416 | -0.762 | 22.188 | 0.454 | 0.948 | -0.256 |
| iCAP 10 | 17.000 | 5.185 | 17.917 | 5.485 | -0.492 | 19.855 | 0.628 | 0.948 | -0.174 |
| iCAP 11 | 8.750 | 4.070 | 8.167 | 5.167 | 0.348 | 17.134 | 0.732 | 0.948 | 0.132 |
| iCAP 12 | 9.250 | 4.291 | 7.500 | 5.018 | 1.054 | 18.246 | 0.306 | 0.948 | 0.388 |
| iCAP 13 | 4.500 | 2.975 | 3.917 | 1.505 | 0.821 | 36.728 | 0.417 | 0.948 | 0.221 |

**Table S3**. Group comparison for the VPT and full-term control groups for the total duration of the identified iCAPs networks

| iCAPs – Total duration | VPT group | | Full-term group | | Group comparison | | | | |
|---|---|---|---|---|---|---|---|---|---|
| | Mean | SD | Mean_FT | SD_FT | t | df | p-value | q-value (fdr) | Effect size (d) |
| iCAP 1 | 44.477 | 8.636 | 49.705 | 8.020 | -1.846 | 22.396 | 0.078 | 0.508 | -0.618 |
| iCAP 2 | 32.072 | 9.216 | 33.036 | 7.019 | -0.361 | 27.211 | 0.721 | 0.781 | -0.112 |
| iCAP 3 | 21.889 | 5.857 | 20.643 | 5.536 | 0.641 | 22.023 | 0.528 | 0.687 | 0.216 |
| iCAP 4 | 19.855 | 4.991 | 17.964 | 6.581 | 0.891 | 16.679 | 0.385 | 0.557 | 0.344 |
| iCAP 5 | 24.793 | 10.264 | 29.196 | 8.310 | -1.427 | 25.624 | 0.166 | 0.525 | -0.452 |
| iCAP 6 | 23.950 | 7.687 | 26.015 | 5.529 | -0.957 | 28.738 | 0.346 | 0.557 | -0.290 |
| iCAP 7 | 23.661 | 6.349 | 20.995 | 6.528 | 1.193 | 20.364 | 0.247 | 0.534 | 0.416 |
| iCAP 8 | 15.501 | 7.707 | 12.746 | 5.295 | 1.305 | 29.972 | 0.202 | 0.525 | 0.388 |
| iCAP 9 | 23.614 | 7.244 | 27.313 | 7.637 | -1.425 | 19.912 | 0.170 | 0.525 | -0.503 |
| iCAP 10 | 17.618 | 6.518 | 17.781 | 5.031 | -0.086 | 26.856 | 0.932 | 0.932 | -0.027 |
| iCAP 11 | 10.408 | 6.744 | 9.193 | 6.657 | 0.527 | 21.136 | 0.604 | 0.713 | 0.181 |
| iCAP 12 | 8.360 | 4.815 | 5.876 | 3.488 | 1.830 | 28.548 | 0.078 | 0.508 | 0.555 |
| iCAP 13 | 4.248 | 3.084 | 3.334 | 2.205 | 1.059 | 28.895 | 0.298 | 0.554 | 0.320 |

**Table S4**. Group comparison for the VPT and full-term control groups for the coupling of the identified iCAPs networks

| Coupling | VPT group | | Full-term group | | Group comparison | | | | |
|---|---|---|---|---|---|---|---|---|---|
| | Mean | SD | Mean | SD | t | df | p-value | q-value (fdr) | Effect size (d) |
| iCAP1 & iCAP2 | 0.116 | 0.068 | 0.121 | 0.054 | -0.241 | 26.174 | 0.812 | 0.972 | -0.076 |
| iCAP1 & iCAP3 | 0.09 | 0.046 | 0.101 | 0.042 | -0.713 | 22.705 | 0.483 | 0.972 | -0.237 |
| iCAP1 & iCAP4 | 0.03 | 0.029 | 0.032 | 0.025 | -0.264 | 24.235 | 0.794 | 0.972 | -0.086 |
| iCAP1 & iCAP5 | 0.064 | 0.025 | 0.067 | 0.039 | -0.228 | 15.255 | 0.823 | 0.972 | -0.093 |
| iCAP1 & iCAP6 | 0.044 | 0.027 | 0.022 | 0.015 | 3.128 | 35.445 | 0.004 | 0.035 | 0.863 |
| iCAP1 & iCAP7 | 0.163 | 0.076 | 0.147 | 0.053 | 0.773 | 29.655 | 0.445 | 0.972 | 0.231 |
| iCAP1 & iCAP8 | 0.075 | 0.046 | 0.065 | 0.04 | 0.748 | 23.552 | 0.462 | 0.972 | 0.245 |
| iCAP1 & iCAP9 | 0.156 | 0.048 | 0.148 | 0.068 | 0.373 | 15.933 | 0.714 | 0.972 | 0.148 |
| iCAP1 & iCAP10 | 0.071 | 0.042 | 0.039 | 0.025 | 2.975 | 33.142 | 0.005 | 0.035 | 0.848 |
| iCAP1 & iCAP11 | 0.05 | 0.048 | 0.05 | 0.042 | -0.034 | 23.643 | 0.973 | 0.973 | -0.011 |
| iCAP1 & iCAP12 | 0.064 | 0.05 | 0.049 | 0.026 | 1.23 | 36.274 | 0.227 | 0.972 | 0.334 |
| iCAP1 & iCAP13 | 0.042 | 0.044 | 0.05 | 0.036 | -0.6 | 25.114 | 0.554 | 0.972 | -0.192 |
| iCAP2 & iCAP3 | 0.101 | 0.059 | 0.103 | 0.054 | -0.065 | 23.057 | 0.949 | 0.973 | -0.021 |
| iCAP2 & iCAP4 | 0.116 | 0.068 | 0.121 | 0.054 | -0.241 | 26.174 | 0.812 | 0.972 | -0.076 |
| iCAP2 & iCAP5 | 0.09 | 0.046 | 0.101 | 0.042 | -0.713 | 22.705 | 0.483 | 0.972 | -0.237 |
| iCAP2 & iCAP6 | 0.03 | 0.029 | 0.032 | 0.025 | -0.264 | 24.235 | 0.794 | 0.972 | -0.086 |
| iCAP2 & iCAP7 | 0.064 | 0.025 | 0.067 | 0.039 | -0.228 | 15.255 | 0.823 | 0.972 | -0.093 |
| iCAP2 & iCAP8 | 0.044 | 0.027 | 0.022 | 0.015 | 3.128 | 35.445 | 0.004 | 0.035 | 0.863 |
| iCAP2 & iCAP9 | 0.163 | 0.076 | 0.147 | 0.053 | 0.773 | 29.655 | 0.445 | 0.972 | 0.231 |
| iCAP2 & iCAP10 | 0.075 | 0.046 | 0.065 | 0.04 | 0.748 | 23.552 | 0.462 | 0.972 | 0.245 |
| iCAP2 & iCAP11 | 0.156 | 0.048 | 0.148 | 0.068 | 0.373 | 15.933 | 0.714 | 0.972 | 0.148 |
| iCAP2 & iCAP12 | 0.071 | 0.042 | 0.039 | 0.025 | 2.975 | 33.142 | 0.005 | 0.035 | 0.848 |
| iCAP2 & iCAP13 | 0.05 | 0.048 | 0.05 | 0.042 | -0.034 | 23.643 | 0.973 | 0.973 | -0.011 |
| iCAP3 & iCAP4 | 0.064 | 0.05 | 0.049 | 0.026 | 1.23 | 36.274 | 0.227 | 0.972 | 0.334 |

| | | | | | | | | | |
|---|---|---|---|---|---|---|---|---|---|
| iCAP3 & iCAP5 | 0.042 | 0.044 | 0.05 | 0.036 | -0.6 | 25.114 | 0.554 | 0.972 | -0.192 |
| iCAP3 & iCAP6 | 0.101 | 0.059 | 0.103 | 0.054 | -0.065 | 23.057 | 0.949 | 0.973 | -0.021 |
| iCAP3 & iCAP7 | 0.116 | 0.068 | 0.121 | 0.054 | -0.241 | 26.174 | 0.812 | 0.972 | -0.076 |
| iCAP3 & iCAP8 | 0.09 | 0.046 | 0.101 | 0.042 | -0.713 | 22.705 | 0.483 | 0.972 | -0.237 |
| iCAP3 & iCAP9 | 0.03 | 0.029 | 0.032 | 0.025 | -0.264 | 24.235 | 0.794 | 0.972 | -0.086 |
| iCAP3 & iCAP10 | 0.064 | 0.025 | 0.067 | 0.039 | -0.228 | 15.255 | 0.823 | 0.972 | -0.093 |
| iCAP3 & iCAP11 | 0.044 | 0.027 | 0.022 | 0.015 | 3.128 | 35.445 | 0.004 | 0.035 | 0.863 |
| iCAP3 & iCAP12 | 0.163 | 0.076 | 0.147 | 0.053 | 0.773 | 29.655 | 0.445 | 0.972 | 0.231 |
| iCAP3 & iCAP13 | 0.075 | 0.046 | 0.065 | 0.04 | 0.748 | 23.552 | 0.462 | 0.972 | 0.245 |
| iCAP4 & iCAP5 | 0.156 | 0.048 | 0.148 | 0.068 | 0.373 | 15.933 | 0.714 | 0.972 | 0.148 |
| iCAP4 & iCAP6 | 0.071 | 0.042 | 0.039 | 0.025 | 2.975 | 33.142 | 0.005 | 0.035 | 0.848 |
| iCAP4 & iCAP7 | 0.05 | 0.048 | 0.05 | 0.042 | -0.034 | 23.643 | 0.973 | 0.973 | -0.011 |
| iCAP4 & iCAP8 | 0.064 | 0.05 | 0.049 | 0.026 | 1.23 | 36.274 | 0.227 | 0.972 | 0.334 |
| iCAP4 & iCAP9 | 0.042 | 0.044 | 0.05 | 0.036 | -0.6 | 25.114 | 0.554 | 0.972 | -0.192 |
| iCAP4 & iCAP10 | 0.101 | 0.059 | 0.103 | 0.054 | -0.065 | 23.057 | 0.949 | 0.973 | -0.021 |
| iCAP4 & iCAP11 | 0.116 | 0.068 | 0.121 | 0.054 | -0.241 | 26.174 | 0.812 | 0.972 | -0.076 |
| iCAP4 & iCAP12 | 0.09 | 0.046 | 0.101 | 0.042 | -0.713 | 22.705 | 0.483 | 0.972 | -0.237 |
| iCAP4 & iCAP13 | 0.03 | 0.029 | 0.032 | 0.025 | -0.264 | 24.235 | 0.794 | 0.972 | -0.086 |
| iCAP5 & iCAP6 | 0.064 | 0.025 | 0.067 | 0.039 | -0.228 | 15.255 | 0.823 | 0.972 | -0.093 |
| iCAP5 & iCAP7 | 0.044 | 0.027 | 0.022 | 0.015 | 3.128 | 35.445 | 0.004 | 0.035 | 0.863 |
| iCAP5 & iCAP8 | 0.163 | 0.076 | 0.147 | 0.053 | 0.773 | 29.655 | 0.445 | 0.972 | 0.231 |
| iCAP5 & iCAP9 | 0.075 | 0.046 | 0.065 | 0.04 | 0.748 | 23.552 | 0.462 | 0.972 | 0.245 |
| iCAP5 & iCAP10 | 0.156 | 0.048 | 0.148 | 0.068 | 0.373 | 15.933 | 0.714 | 0.972 | 0.148 |
| iCAP5 & iCAP11 | 0.071 | 0.042 | 0.039 | 0.025 | 2.975 | 33.142 | 0.005 | 0.035 | 0.848 |
| iCAP5 & iCAP12 | 0.05 | 0.048 | 0.05 | 0.042 | -0.034 | 23.643 | 0.973 | 0.973 | -0.011 |
| iCAP5 & iCAP13 | 0.064 | 0.05 | 0.049 | 0.026 | 1.23 | 36.274 | 0.227 | 0.972 | 0.334 |
| iCAP6 & iCAP7 | 0.042 | 0.044 | 0.05 | 0.036 | -0.6 | 25.114 | 0.554 | 0.972 | -0.192 |
| iCAP6 & iCAP8 | 0.101 | 0.059 | 0.103 | 0.054 | -0.065 | 23.057 | 0.949 | 0.973 | -0.021 |

| | | | | | | | | | |
|---|---|---|---|---|---|---|---|---|---|
| iCAP6 & iCAP9 | 0.116 | 0.068 | 0.121 | 0.054 | -0.241 | 26.174 | 0.812 | 0.972 | -0.076 |
| iCAP6 & iCAP10 | 0.09 | 0.046 | 0.101 | 0.042 | -0.713 | 22.705 | 0.483 | 0.972 | -0.237 |
| iCAP6 & iCAP11 | 0.03 | 0.029 | 0.032 | 0.025 | -0.264 | 24.235 | 0.794 | 0.972 | -0.086 |
| iCAP6 & iCAP12 | 0.064 | 0.025 | 0.067 | 0.039 | -0.228 | 15.255 | 0.823 | 0.972 | -0.093 |
| iCAP6 & iCAP13 | 0.044 | 0.027 | 0.022 | 0.015 | 3.128 | 35.445 | 0.004 | 0.035 | 0.863 |
| iCAP7 & iCAP8 | 0.163 | 0.076 | 0.147 | 0.053 | 0.773 | 29.655 | 0.445 | 0.972 | 0.231 |
| iCAP7 & iCAP9 | 0.075 | 0.046 | 0.065 | 0.04 | 0.748 | 23.552 | 0.462 | 0.972 | 0.245 |
| iCAP7 & iCAP10 | 0.156 | 0.048 | 0.148 | 0.068 | 0.373 | 15.933 | 0.714 | 0.972 | 0.148 |
| iCAP7 & iCAP11 | 0.071 | 0.042 | 0.039 | 0.025 | 2.975 | 33.142 | 0.005 | 0.035 | 0.848 |
| iCAP7 & iCAP12 | 0.05 | 0.048 | 0.05 | 0.042 | -0.034 | 23.643 | 0.973 | 0.973 | -0.011 |
| iCAP7 & iCAP13 | 0.064 | 0.05 | 0.049 | 0.026 | 1.23 | 36.274 | 0.227 | 0.972 | 0.334 |
| iCAP8 & iCAP9 | 0.042 | 0.044 | 0.05 | 0.036 | -0.6 | 25.114 | 0.554 | 0.972 | -0.192 |
| iCAP8 & iCAP10 | 0.101 | 0.059 | 0.103 | 0.054 | -0.065 | 23.057 | 0.949 | 0.973 | -0.021 |
| iCAP8 & iCAP11 | 0.116 | 0.068 | 0.121 | 0.054 | -0.241 | 26.174 | 0.812 | 0.972 | -0.076 |
| iCAP8 & iCAP12 | 0.09 | 0.046 | 0.101 | 0.042 | -0.713 | 22.705 | 0.483 | 0.972 | -0.237 |
| iCAP8 & iCAP13 | 0.03 | 0.029 | 0.032 | 0.025 | -0.264 | 24.235 | 0.794 | 0.972 | -0.086 |
| iCAP9 & iCAP10 | 0.064 | 0.025 | 0.067 | 0.039 | -0.228 | 15.255 | 0.823 | 0.972 | -0.093 |
| iCAP9 & iCAP11 | 0.044 | 0.027 | 0.022 | 0.015 | 3.128 | 35.445 | 0.004 | 0.035 | 0.863 |
| iCAP9 & iCAP12 | 0.163 | 0.076 | 0.147 | 0.053 | 0.773 | 29.655 | 0.445 | 0.972 | 0.231 |
| iCAP9 & iCAP13 | 0.075 | 0.046 | 0.065 | 0.04 | 0.748 | 23.552 | 0.462 | 0.972 | 0.245 |
| iCAP10 & iCAP11 | 0.156 | 0.048 | 0.148 | 0.068 | 0.373 | 15.933 | 0.714 | 0.972 | 0.148 |
| iCAP10 & iCAP12 | 0.071 | 0.042 | 0.039 | 0.025 | 2.975 | 33.142 | 0.005 | 0.035 | 0.848 |
| iCAP10 & iCAP13 | 0.05 | 0.048 | 0.05 | 0.042 | -0.034 | 23.643 | 0.973 | 0.973 | -0.011 |
| iCAP11 & iCAP12 | 0.064 | 0.05 | 0.049 | 0.026 | 1.23 | 36.274 | 0.227 | 0.972 | 0.334 |
| iCAP11 & iCAP13 | 0.042 | 0.044 | 0.05 | 0.036 | -0.6 | 25.114 | 0.554 | 0.972 | -0.192 |
| iCAP12 & iCAP13 | 0.101 | 0.059 | 0.103 | 0.054 | -0.065 | 23.057 | 0.949 | 0.973 | -0.021 |

**Table S5**. Group comparison for the VPT and full-term control groups for the anti-coupling of the identified iCAPs networks

| Anti-coupling | VPT group | | Full-term group | | Group comparison | | | | |
|---|---|---|---|---|---|---|---|---|---|
| | Mean | SD | Mean | SD | t | df | p-value | q-value (fdr) | Effect size (d) |
| iCAP1 & iCAP2 | 0.161 | 0.069 | 0.155 | 0.068 | 0.260 | 21.047 | 0.797 | 0.864 | 0.090 |
| iCAP1 & iCAP3 | 0.118 | 0.041 | 0.124 | 0.065 | -0.272 | 14.907 | 0.789 | 0.864 | -0.112 |
| iCAP1 & iCAP4 | 0.236 | 0.071 | 0.215 | 0.117 | 0.567 | 14.617 | 0.579 | 0.837 | 0.237 |
| iCAP1 & iCAP5 | 0.176 | 0.086 | 0.200 | 0.063 | -1.007 | 28.535 | 0.322 | 0.639 | -0.306 |
| iCAP1 & iCAP6 | 0.239 | 0.081 | 0.305 | 0.090 | -2.196 | 19.025 | 0.041 | 0.333 | -0.791 |
| iCAP1 & iCAP7 | 0.078 | 0.066 | 0.063 | 0.037 | 0.955 | 34.873 | 0.346 | 0.639 | 0.266 |
| iCAP1 & iCAP8 | 0.084 | 0.048 | 0.083 | 0.047 | 0.031 | 21.265 | 0.975 | 0.975 | 0.011 |
| iCAP1 & iCAP9 | 0.075 | 0.044 | 0.087 | 0.041 | -0.871 | 22.331 | 0.393 | 0.639 | -0.292 |
| iCAP1 & iCAP10 | 0.117 | 0.071 | 0.162 | 0.059 | -2.049 | 24.785 | 0.051 | 0.333 | -0.658 |
| iCAP1 & iCAP11 | 0.081 | 0.067 | 0.062 | 0.046 | 0.998 | 29.933 | 0.326 | 0.639 | 0.297 |
| iCAP1 & iCAP12 | 0.040 | 0.038 | 0.027 | 0.027 | 1.230 | 29.347 | 0.228 | 0.639 | 0.369 |
| iCAP1 & iCAP13 | 0.020 | 0.023 | 0.010 | 0.015 | 1.684 | 31.680 | 0.102 | 0.442 | 0.490 |
| iCAP2 & iCAP3 | 0.111 | 0.052 | 0.121 | 0.100 | -0.337 | 13.591 | 0.741 | 0.864 | -0.148 |
| iCAP2 & iCAP4 | 0.161 | 0.069 | 0.155 | 0.068 | 0.260 | 21.047 | 0.797 | 0.864 | 0.090 |
| iCAP2 & iCAP5 | 0.118 | 0.041 | 0.124 | 0.065 | -0.272 | 14.907 | 0.789 | 0.864 | -0.112 |
| iCAP2 & iCAP6 | 0.236 | 0.071 | 0.215 | 0.117 | 0.567 | 14.617 | 0.579 | 0.837 | 0.237 |
| iCAP2 & iCAP7 | 0.176 | 0.086 | 0.200 | 0.063 | -1.007 | 28.535 | 0.322 | 0.639 | -0.306 |
| iCAP2 & iCAP8 | 0.239 | 0.081 | 0.305 | 0.090 | -2.196 | 19.025 | 0.041 | 0.333 | -0.791 |
| iCAP2 & iCAP9 | 0.078 | 0.066 | 0.063 | 0.037 | 0.955 | 34.873 | 0.346 | 0.639 | 0.266 |
| iCAP2 & iCAP10 | 0.084 | 0.048 | 0.083 | 0.047 | 0.031 | 21.265 | 0.975 | 0.975 | 0.011 |
| iCAP2 & iCAP11 | 0.075 | 0.044 | 0.087 | 0.041 | -0.871 | 22.331 | 0.393 | 0.639 | -0.292 |
| iCAP2 & iCAP12 | 0.117 | 0.071 | 0.162 | 0.059 | -2.049 | 24.785 | 0.051 | 0.333 | -0.658 |
| iCAP2 & iCAP13 | 0.081 | 0.067 | 0.062 | 0.046 | 0.998 | 29.933 | 0.326 | 0.639 | 0.297 |
| iCAP3 & iCAP4 | 0.040 | 0.038 | 0.027 | 0.027 | 1.230 | 29.347 | 0.228 | 0.639 | 0.369 |

| | | | | | | | | | |
|---|---|---|---|---|---|---|---|---|---|
| iCAP3 & iCAP5 | 0.020 | 0.023 | 0.010 | 0.015 | 1.684 | 31.680 | 0.102 | 0.442 | 0.490 |
| iCAP3 & iCAP6 | 0.111 | 0.052 | 0.121 | 0.100 | -0.337 | 13.591 | 0.741 | 0.864 | -0.148 |
| iCAP3 & iCAP7 | 0.161 | 0.069 | 0.155 | 0.068 | 0.260 | 21.047 | 0.797 | 0.864 | 0.090 |
| iCAP3 & iCAP8 | 0.118 | 0.041 | 0.124 | 0.065 | -0.272 | 14.907 | 0.789 | 0.864 | -0.112 |
| iCAP3 & iCAP9 | 0.236 | 0.071 | 0.215 | 0.117 | 0.567 | 14.617 | 0.579 | 0.837 | 0.237 |
| iCAP3 & iCAP10 | 0.176 | 0.086 | 0.200 | 0.063 | -1.007 | 28.535 | 0.322 | 0.639 | -0.306 |
| iCAP3 & iCAP11 | 0.239 | 0.081 | 0.305 | 0.090 | -2.196 | 19.025 | 0.041 | 0.333 | -0.791 |
| iCAP3 & iCAP12 | 0.078 | 0.066 | 0.063 | 0.037 | 0.955 | 34.873 | 0.346 | 0.639 | 0.266 |
| iCAP3 & iCAP13 | 0.084 | 0.048 | 0.083 | 0.047 | 0.031 | 21.265 | 0.975 | 0.975 | 0.011 |
| iCAP4 & iCAP5 | 0.075 | 0.044 | 0.087 | 0.041 | -0.871 | 22.331 | 0.393 | 0.639 | -0.292 |
| iCAP4 & iCAP6 | 0.117 | 0.071 | 0.162 | 0.059 | -2.049 | 24.785 | 0.051 | 0.333 | -0.658 |
| iCAP4 & iCAP7 | 0.081 | 0.067 | 0.062 | 0.046 | 0.998 | 29.933 | 0.326 | 0.639 | 0.297 |
| iCAP4 & iCAP8 | 0.040 | 0.038 | 0.027 | 0.027 | 1.230 | 29.347 | 0.228 | 0.639 | 0.369 |
| iCAP4 & iCAP9 | 0.020 | 0.023 | 0.010 | 0.015 | 1.684 | 31.680 | 0.102 | 0.442 | 0.490 |
| iCAP4 & iCAP10 | 0.111 | 0.052 | 0.121 | 0.100 | -0.337 | 13.591 | 0.741 | 0.864 | -0.148 |
| iCAP4 & iCAP11 | 0.161 | 0.069 | 0.155 | 0.068 | 0.260 | 21.047 | 0.797 | 0.864 | 0.090 |
| iCAP4 & iCAP12 | 0.118 | 0.041 | 0.124 | 0.065 | -0.272 | 14.907 | 0.789 | 0.864 | -0.112 |
| iCAP4 & iCAP13 | 0.236 | 0.071 | 0.215 | 0.117 | 0.567 | 14.617 | 0.579 | 0.837 | 0.237 |
| iCAP5 & iCAP6 | 0.176 | 0.086 | 0.200 | 0.063 | -1.007 | 28.535 | 0.322 | 0.639 | -0.306 |
| iCAP5 & iCAP7 | 0.239 | 0.081 | 0.305 | 0.090 | -2.196 | 19.025 | 0.041 | 0.333 | -0.791 |
| iCAP5 & iCAP8 | 0.078 | 0.066 | 0.063 | 0.037 | 0.955 | 34.873 | 0.346 | 0.639 | 0.266 |
| iCAP5 & iCAP9 | 0.084 | 0.048 | 0.083 | 0.047 | 0.031 | 21.265 | 0.975 | 0.975 | 0.011 |
| iCAP5 & iCAP10 | 0.075 | 0.044 | 0.087 | 0.041 | -0.871 | 22.331 | 0.393 | 0.639 | -0.292 |
| iCAP5 & iCAP11 | 0.117 | 0.071 | 0.162 | 0.059 | -2.049 | 24.785 | 0.051 | 0.333 | -0.658 |
| iCAP5 & iCAP12 | 0.081 | 0.067 | 0.062 | 0.046 | 0.998 | 29.933 | 0.326 | 0.639 | 0.297 |
| iCAP5 & iCAP13 | 0.040 | 0.038 | 0.027 | 0.027 | 1.230 | 29.347 | 0.228 | 0.639 | 0.369 |
| iCAP6 & iCAP7 | 0.020 | 0.023 | 0.010 | 0.015 | 1.684 | 31.680 | 0.102 | 0.442 | 0.490 |
| iCAP6 & iCAP8 | 0.111 | 0.052 | 0.121 | 0.100 | -0.337 | 13.591 | 0.741 | 0.864 | -0.148 |

| | | | | | | | | | |
|---|---|---|---|---|---|---|---|---|---|
| iCAP6 & iCAP9 | 0.161 | 0.069 | 0.155 | 0.068 | 0.260 | 21.047 | 0.797 | 0.864 | 0.090 |
| iCAP6 & iCAP10 | 0.118 | 0.041 | 0.124 | 0.065 | -0.272 | 14.907 | 0.789 | 0.864 | -0.112 |
| iCAP6 & iCAP11 | 0.236 | 0.071 | 0.215 | 0.117 | 0.567 | 14.617 | 0.579 | 0.837 | 0.237 |
| iCAP6 & iCAP12 | 0.176 | 0.086 | 0.200 | 0.063 | -1.007 | 28.535 | 0.322 | 0.639 | -0.306 |
| iCAP6 & iCAP13 | 0.239 | 0.081 | 0.305 | 0.090 | -2.196 | 19.025 | 0.041 | 0.333 | -0.791 |
| iCAP7 & iCAP8 | 0.078 | 0.066 | 0.063 | 0.037 | 0.955 | 34.873 | 0.346 | 0.639 | 0.266 |
| iCAP7 & iCAP9 | 0.084 | 0.048 | 0.083 | 0.047 | 0.031 | 21.265 | 0.975 | 0.975 | 0.011 |
| iCAP7 & iCAP10 | 0.075 | 0.044 | 0.087 | 0.041 | -0.871 | 22.331 | 0.393 | 0.639 | -0.292 |
| iCAP7 & iCAP11 | 0.117 | 0.071 | 0.162 | 0.059 | -2.049 | 24.785 | 0.051 | 0.333 | -0.658 |
| iCAP7 & iCAP12 | 0.081 | 0.067 | 0.062 | 0.046 | 0.998 | 29.933 | 0.326 | 0.639 | 0.297 |
| iCAP7 & iCAP13 | 0.040 | 0.038 | 0.027 | 0.027 | 1.230 | 29.347 | 0.228 | 0.639 | 0.369 |
| iCAP8 & iCAP9 | 0.020 | 0.023 | 0.010 | 0.015 | 1.684 | 31.680 | 0.102 | 0.442 | 0.490 |
| iCAP8 & iCAP10 | 0.111 | 0.052 | 0.121 | 0.100 | -0.337 | 13.591 | 0.741 | 0.864 | -0.148 |
| iCAP8 & iCAP11 | 0.161 | 0.069 | 0.155 | 0.068 | 0.260 | 21.047 | 0.797 | 0.864 | 0.090 |
| iCAP8 & iCAP12 | 0.118 | 0.041 | 0.124 | 0.065 | -0.272 | 14.907 | 0.789 | 0.864 | -0.112 |
| iCAP8 & iCAP13 | 0.236 | 0.071 | 0.215 | 0.117 | 0.567 | 14.617 | 0.579 | 0.837 | 0.237 |
| iCAP9 & iCAP10 | 0.176 | 0.086 | 0.200 | 0.063 | -1.007 | 28.535 | 0.322 | 0.639 | -0.306 |
| iCAP9 & iCAP11 | 0.239 | 0.081 | 0.305 | 0.090 | -2.196 | 19.025 | 0.041 | 0.333 | -0.791 |
| iCAP9 & iCAP12 | 0.078 | 0.066 | 0.063 | 0.037 | 0.955 | 34.873 | 0.346 | 0.639 | 0.266 |
| iCAP9 & iCAP13 | 0.084 | 0.048 | 0.083 | 0.047 | 0.031 | 21.265 | 0.975 | 0.975 | 0.011 |
| iCAP10 & iCAP11 | 0.075 | 0.044 | 0.087 | 0.041 | -0.871 | 22.331 | 0.393 | 0.639 | -0.292 |
| iCAP10 & iCAP12 | 0.117 | 0.071 | 0.162 | 0.059 | -2.049 | 24.785 | 0.051 | 0.333 | -0.658 |
| iCAP10 & iCAP13 | 0.081 | 0.067 | 0.062 | 0.046 | 0.998 | 29.933 | 0.326 | 0.639 | 0.297 |
| iCAP11 & iCAP12 | 0.040 | 0.038 | 0.027 | 0.027 | 1.230 | 29.347 | 0.228 | 0.639 | 0.369 |
| iCAP11 & iCAP13 | 0.020 | 0.023 | 0.010 | 0.015 | 1.684 | 31.680 | 0.102 | 0.442 | 0.490 |
| iCAP12 & iCAP13 | 0.111 | 0.052 | 0.121 | 0.100 | -0.337 | 13.591 | 0.741 | 0.864 | -0.148 |

**Table S6**. Group comparison for the VPT and full-term control groups for the Amplitude-Laterality Index (A-LI) of the identified iCAPs networks

| Amplitude-LI | VPT group | | Full-term group | | Group comparison | | | | |
|---|---|---|---|---|---|---|---|---|---|
| | Mean | SD | Mean_FT | SD_FT | t | df | p-value | q-value (fdr) | Effect size (d) |
| iCAP 1 | 0.027 | 0.034 | 0.030 | 0.046 | -0.203 | 16.268 | 0.842 | 0.984 | -0.079 |
| iCAP 2 | 0.043 | 0.032 | 0.049 | 0.036 | -0.514 | 18.613 | 0.613 | 0.984 | -0.187 |
| iCAP 3 | 0.024 | 0.056 | 0.029 | 0.061 | -0.239 | 19.421 | 0.814 | 0.984 | -0.085 |
| iCAP 4 | 0.024 | 0.061 | 0.014 | 0.062 | 0.471 | 20.509 | 0.642 | 0.984 | 0.164 |
| iCAP 5 | 0.009 | 0.038 | 0.008 | 0.047 | 0.020 | 17.263 | 0.984 | 0.984 | 0.008 |
| iCAP 6 | 0.063 | 0.081 | 0.024 | 0.040 | 2.047 | 37.099 | 0.048 | 0.311 | 0.547 |
| iCAP 7 | 0.395 | 0.124 | 0.423 | 0.098 | -0.739 | 26.202 | 0.466 | 0.984 | -0.232 |
| iCAP 8 | 0.024 | 0.045 | 0.009 | 0.055 | 0.834 | 17.721 | 0.416 | 0.984 | 0.311 |
| iCAP 9 | -0.393 | 0.118 | -0.390 | 0.110 | -0.085 | 22.175 | 0.933 | 0.984 | -0.029 |
| iCAP 10 | 0.041 | 0.085 | 0.026 | 0.078 | 0.555 | 22.577 | 0.584 | 0.984 | 0.185 |
| iCAP 11 | 0.029 | 0.047 | 0.040 | 0.049 | -0.639 | 19.917 | 0.530 | 0.984 | -0.225 |
| iCAP 12 | 0.019 | 0.048 | 0.064 | 0.039 | -3.049 | 25.465 | 0.005 | 0.069 | -0.968 |
| iCAP 13 | 0.030 | 0.076 | 0.022 | 0.090 | 0.263 | 18.077 | 0.795 | 0.984 | 0.097 |

**Table S7**. *Associations between socio-emotional outcomes and occurrence of the 13 iCAPs in the very preterm (VPT) and full-term (FT) groups based on the PLSC analysis, corresponding to Figure 1. The table shows mean saliences, bootstrap-estimated standard deviations and bootstrap ratio Z-scores for socio-emotional and iCAPs occurrence measures for the significant latent component 1.*

| ***Salience type: Socio-emotional measures*** | ***Very preterm group*** | | ***Full-term control group*** | |
|---|---|---|---|---|
| | Mean salience (bootstrap estimated standard deviation) | Bootstrap ratio Z-scores | Mean salience (bootstrap estimated standard deviation) | Bootstrap ratio Z-scores |
| *Affect recognition* | *0.05 (0.11)* | *0.80* | *0.26 (0.12)* | *4.97* |
| *Theory of mind* | *-0.11 (0.15)* | *-0.82* | *0.14 (0.13)* | *4.42* |
| *Internalised problems* | *-0.03 (0.15)* | *-0.47* | *-0.02 (0.12)* | *-2.10* |
| *Emotional control* | *0.35 (0.17)* | *2.47* | *0.09 (0.15)* | *-1.65* |

| ***Coupling duration measures*** | ***For both the very preterm and full-term groups*** | |
|---|---|---|
| | Mean salience (bootstrap estimated standard deviation) | Bootstrap ratio Z-scores |
| *iCAP 1* | *0.02 (0.1)* | *-1.55* |
| *iCAP 2* | *-0.13 (0.14)* | *-1.37* |
| *iCAP 3* | *-0.2 (0.11)* | *-3.74* |
| *iCAP 4* | *-0.18 (0.12)* | *-2.46* |
| *iCAP 5* | *-0.01 (0.12)* | *-1.61* |
| *iCAP 6* | *-0.07 (0.11)* | *-3.20* |
| *iCAP 7* | *-0.1 (0.1)* | *-1.71* |
| *iCAP 8* | *-0.07 (0.12)* | *-2.56* |
| *iCAP 9* | *-0.14 (0.1)* | *-3.54* |
| *iCAP 10* | *0.01 (0.1)* | *-3.28* |
| *iCAP 11* | *0.2 (0.11)* | *0.01* |
| *iCAP 12* | *-0.14 (0.14)* | *-2.02* |
| *iCAP 13* | *0.12 (0.12)* | *2.33* |

**Table S8**. *Associations between socio-emotional outcomes and coupling duration of the 13 pair of iCAPs in the very preterm (VPT) and full-term (FT) groups based on the PLSC analysis, corresponding to Figure 2. The table shows mean saliences, bootstrap-estimated standard deviations and bootstrap ratio Z-scores for socio-emotional and iCAPs coupling measures for the significant latent component 1.*

| ***Salience type: Socio-emotional measures*** | ***Very preterm group*** | | ***Full-term control group*** | |
|---|---|---|---|---|
| | Mean salience (bootstrap estimated standard deviation) | Bootstrap ratio Z-scores | Mean salience (bootstrap estimated standard deviation) | Bootstrap ratio Z-scores |
| *Affect recognition* | *-0.15 (0.07)* | *-2.63* | *-0.18 (0.07)* | *-2.55* |
| *Theory of mind* | *0.07 (0.05)* | *1.48* | *0.01 (0.08)* | *-1.14* |
| *Internalised problems* | *-0.3 (0.06)* | *-5.55* | *-0.08 (0.07)* | *-1.12* |
| *Emotional control* | *-0.82 (0.06)* | *-14.07* | *-0.15 (0.08)* | *1.08* |

| ***Coupling duration measures*** | ***For both the very preterm and full-term groups*** | |
|---|---|---|
| | Mean salience (bootstrap estimated standard deviation) | Bootstrap ratio Z-scores |
| *iCAP1 & iCAP2* | *-1.34* | *-0.05 (0.05)* |
| *iCAP1 & iCAP3* | *5.54* | *0.16 (0.04)* |
| *iCAP1 & iCAP4* | *0.79* | *0.04 (0.05)* |
| *iCAP1 & iCAP5* | *0.30* | *0.02 (0.06)* |
| *iCAP1 & iCAP6* | *-6.17* | *-0.19 (0.04)* |
| *iCAP1 & iCAP7* | *-0.19* | *-0.02 (0.05)* |
| *iCAP1 & iCAP8* | *-0.96* | *-0.01 (0.05)* |
| *iCAP1 & iCAP9* | *-1.43* | *-0.06 (0.04)* |
| *iCAP1 & iCAP10* | *0.52* | *0.02 (0.05)* |
| *iCAP1 & iCAP11* | *-1.02* | *-0.05 (0.08)* |
| *iCAP1 & iCAP12* | *-0.72* | *-0.04 (0.05)* |
| *iCAP1 & iCAP13* | *-2.20* | *-0.07 (0.04)* |
| *iCAP2 & iCAP3* | *-3.12* | *-0.12 (0.05)* |
| *iCAP2 & iCAP4* | *0.23* | *0.02 (0.04)* |
| *iCAP2 & iCAP5* | *0.27* | *0.03 (0.04)* |
| *iCAP2 & iCAP6* | *-0.69* | *-0.04 (0.06)* |
| *iCAP2 & iCAP7* | *-0.32* | *-0.02 (0.06)* |
| *iCAP2 & iCAP8* | *-2.15* | *-0.11 (0.06)* |
| *iCAP2 & iCAP9* | *-4.16* | *-0.16 (0.04)* |

| | | |
|---|---|---|
| *iCAP2 & iCAP10* | *0.49* | *0.01 (0.05)* |
| *iCAP2 & iCAP11* | *-1.22* | *-0.08 (0.07)* |
| *iCAP2 & iCAP12* | *-3.02* | *-0.15 (0.05)* |
| *iCAP2 & iCAP13* | *-3.21* | *-0.11 (0.04)* |
| *iCAP3 & iCAP4* | *-1.06* | *-0.04 (0.06)* |
| *iCAP3 & iCAP5* | *-2.42* | *-0.1 (0.05)* |
| *iCAP3 & iCAP6* | *-1.15* | *-0.05 (0.07)* |
| *iCAP3 & iCAP7* | *1.63* | *0.07 (0.05)* |
| *iCAP3 & iCAP8* | *-1.92* | *-0.08 (0.05)* |
| *iCAP3 & iCAP9* | *1.65* | *0.07 (0.06)* |
| *iCAP3 & iCAP10* | *-0.98* | *-0.04 (0.07)* |
| *iCAP3 & iCAP11* | *-0.36* | *0.01 (0.07)* |
| *iCAP3 & iCAP12* | *-2.72* | *-0.13 (0.05)* |
| *iCAP3 & iCAP13* | *-2.98* | *-0.12 (0.04)* |
| *iCAP4 & iCAP5* | *-1.44* | *-0.05 (0.06)* |
| *iCAP4 & iCAP6* | *4.98* | *0.19 (0.04)* |
| *iCAP4 & iCAP7* | *-0.52* | *-0.01 (0.06)* |
| *iCAP4 & iCAP8* | *-1.18* | *-0.06 (0.05)* |
| *iCAP4 & iCAP9* | *0.06* | *0 (0.04)* |
| *iCAP4 & iCAP10* | *-3.83* | *-0.17 (0.06)* |
| *iCAP4 & iCAP11* | *-0.05* | *-0.01 (0.06)* |
| *iCAP4 & iCAP12* | *-2.07* | *-0.09 (0.06)* |
| *iCAP4 & iCAP13* | *-2.10* | *-0.08 (0.04)* |
| *iCAP5 & iCAP6* | *1.75* | *0.07 (0.06)* |
| *iCAP5 & iCAP7* | *-0.09* | *-0.03 (0.07)* |
| *iCAP5 & iCAP8* | *-9.46* | *-0.21 (0.03)* |
| *iCAP5 & iCAP9* | *-2.59* | *-0.15 (0.06)* |
| *iCAP5 & iCAP10* | *-3.98* | *-0.14 (0.05)* |
| *iCAP5 & iCAP11* | *-0.01* | *0.01 (0.05)* |
| *iCAP5 & iCAP12* | *0.32* | *0.02 (0.05)* |
| *iCAP5 & iCAP13* | *-2.36* | *-0.09 (0.05)* |
| *iCAP6 & iCAP7* | *-0.14* | *-0.02 (0.08)* |
| *iCAP6 & iCAP8* | *-1.61* | *-0.06 (0.06)* |
| *iCAP6 & iCAP9* | *-4.83* | *-0.21 (0.05)* |
| *iCAP6 & iCAP10* | *0.43* | *0.02 (0.07)* |

| | | |
|---|---|---|
| *iCAP6 & iCAP11* | *-0.72* | *-0.04 (0.06)* |
| *iCAP6 & iCAP12* | *-0.34* | *-0.01 (0.04)* |
| *iCAP6 & iCAP13* | *-3.51* | *-0.15 (0.05)* |
| *iCAP7 & iCAP8* | *0.04* | *0.01 (0.07)* |
| *iCAP7 & iCAP9* | *-1.67* | *-0.08 (0.05)* |
| *iCAP7 & iCAP10* | *-0.09* | *0 (0.06)* |
| *iCAP7 & iCAP11* | *-1.88* | *-0.07 (0.06)* |
| *iCAP7 & iCAP12* | *-2.84* | *-0.12 (0.06)* |
| *iCAP7 & iCAP13* | *0.19* | *0 (0.05)* |
| *iCAP8 & iCAP9* | *-1.71* | *-0.07 (0.06)* |
| *iCAP8 & iCAP10* | *-3.07* | *-0.12 (0.05)* |
| *iCAP8 & iCAP11* | *-0.20* | *0.01 (0.06)* |
| *iCAP8 & iCAP12* | *3.50* | *0.13 (0.04)* |
| *iCAP8 & iCAP13* | *-2.21* | *-0.1 (0.06)* |
| *iCAP9 & iCAP10* | *4.55* | *0.14 (0.04)* |
| *iCAP9 & iCAP11* | *-2.64* | *-0.09 (0.05)* |
| *iCAP9 & iCAP12* | *-3.24* | *-0.12 (0.04)* |
| *iCAP9 & iCAP13* | *-4.14* | *-0.12 (0.05)* |
| *iCAP10 & iCAP11* | *-1.77* | *-0.07 (0.05)* |
| *iCAP10 & iCAP12* | *-1.07* | *-0.05 (0.06)* |
| *iCAP10 & iCAP13* | *-0.67* | *-0.03 (0.04)* |
| *iCAP11 & iCAP12* | *-2.41* | *-0.09 (0.04)* |
| *iCAP11 & iCAP13* | *-4.10* | *-0.13 (0.04)* |
| *iCAP12 & iCAP13* | *-0.56* | *-0.02 (0.07)* |
